# Long-Term Impact of Cumulative Hyperglycaemia on DNA Methylation and its Role in Diabetic Kidney Disease

**DOI:** 10.64898/2026.08.31.26361614

**Authors:** Xiaoqi Luo, Anna Syreeni, Claire Hill, Laura J. Smyth, Emma H. Dahlström, Stefan Mutter, Zhuo Chen, Rama Natarajan, DCCT/EDIC Study Group, Siyu Pan, Andrew Parton, Heather Jackson, Gareth McKay, Katalin Susztak, Joel N. Hirschhorn, Jose C. Florez, GENIE Consortium, Alexander P. Maxwell, Per-Henrik Groop, Amy Jayne McKnight, Niina Sandholm, FinnDiane Study Group

## Abstract

Hyperglycaemia is a hallmark of diabetes and a major risk factor for diabetic kidney disease (DKD). However, the molecular consequences of long-term cumulative hyperglycaemia (CH) remain unclear. As a stable epigenetic modification, DNA methylation may capture past glycaemic exposure. Here, we assessed CH-associated DNA methylation in 1,245 participants with type 1 diabetes (T1D) from Finland and the United Kingdom-Republic of Ireland cohorts. We identified 17 CH-associated CpGs, with the strongest association at cg19693031 (*TXNIP*). Longitudinal analyses demonstrate that these CH-associated DNA methylation levels remain stable despite short-term glycaemic fluctuations, suggesting lasting epigenetic imprints of earlier metabolic control. Integrative analyses combining genomic, epigenetic, and proteomic data characterized these CpGs and potential target proteins. Mendelian randomization suggested a causal association between cg20853880 (*KLF11*) and DKD, supported by chromatin accessibility and kidney *KLF11* expression. Our findings suggest that epigenetic changes contribute to metabolic memory and may mediate the effects of hyperglycaemia on DKD.

## Introduction

Diabetes is a major health challenge affecting approximately 589 million individuals globally^1^, with type 1 diabetes (T1D) accounting for 9.5 million cases^2^. A majority of individuals with diabetes will develop some micro-and macrovascular complications. Hyperglycaemia is the main driver of diabetic complications, including diabetic kidney disease (DKD) which is present in 30-40% of persons with T1D^3^. Previous studies from the Diabetes Control and Complications Trial (DCCT) and the Epidemiology of Diabetes Interventions and Complications (EDIC) follow-up study observed that earlier glycaemic exposure has persistent effect on future risk of diabetic complications^4,5^. This phenomenon is known as metabolic memory. Epigenetic modifications are proposed to contribute to metabolic memory, as the epigenome-modifying enzymes activity depends on intermediary metabolism substrates such as acetyl-CoA and therefore, is highly responsive to intracellular metabolite fluctuations^6^. Moreover, while DNA methylation changes are reversible, they may also persist for long periods under disease conditions. For instance, previous studies on diabetic foot ulcers identified sustained DNA methylation levels in fibroblasts after prolonged passage in normoglycemic conditions^7^.

Mediation and Mendelian Randomisation (MR) analysis has been widely used to identify potential causal pathways connecting exposures and outcomes. Previous mediation and MR studies on mean-HbA1c implied that HbA1c-associated CpGs play a potential mediating and causal role in diabetic complications^8^. Accelerated kidney ageing has also been suggested as a risk factor in DKD pathogenesis^9^, and recent studies have identified significant associations between epigenetic age acceleration (EAA) and kidney function decline^10^. However, the association between EAA and cumulative hyperglycaemia (CH) remains poorly understood.

Previous investigations have already demonstrated that hyperglycaemia exposure can result in long-lasting effects^8^. Epigenome-wide association studies (EWAS) on hyperglycaemia in T1D have mainly assessed glycaemic exposure at a single time point or pooled as mean HbA1c^8,11^, which reflects either baseline or long-term average glycaemic control. Here, we extend previous work by focusing on how cumulative burden of hyperglycaemia affects epigenetics and may ultimately contribute to DKD progression. In this study, we report findings from an EWAS meta-analysis of CH in 1245 adult participants with long duration of T1D, with expanded CpG coverage and a 2.5-fold increase in sample size compared with previous studies^8,11^. Furthermore, we assess the role of DNA methylation in mediating the link between CH and DKD and identify causal associations between hyperglycaemia-associated DNA methylation changes and DKD (**Fig. 1**). Altogether, the current study proposes novel mechanisms underlying the long-term impact of CH on DNA methylation and gene regulation.

**Figure 1.**
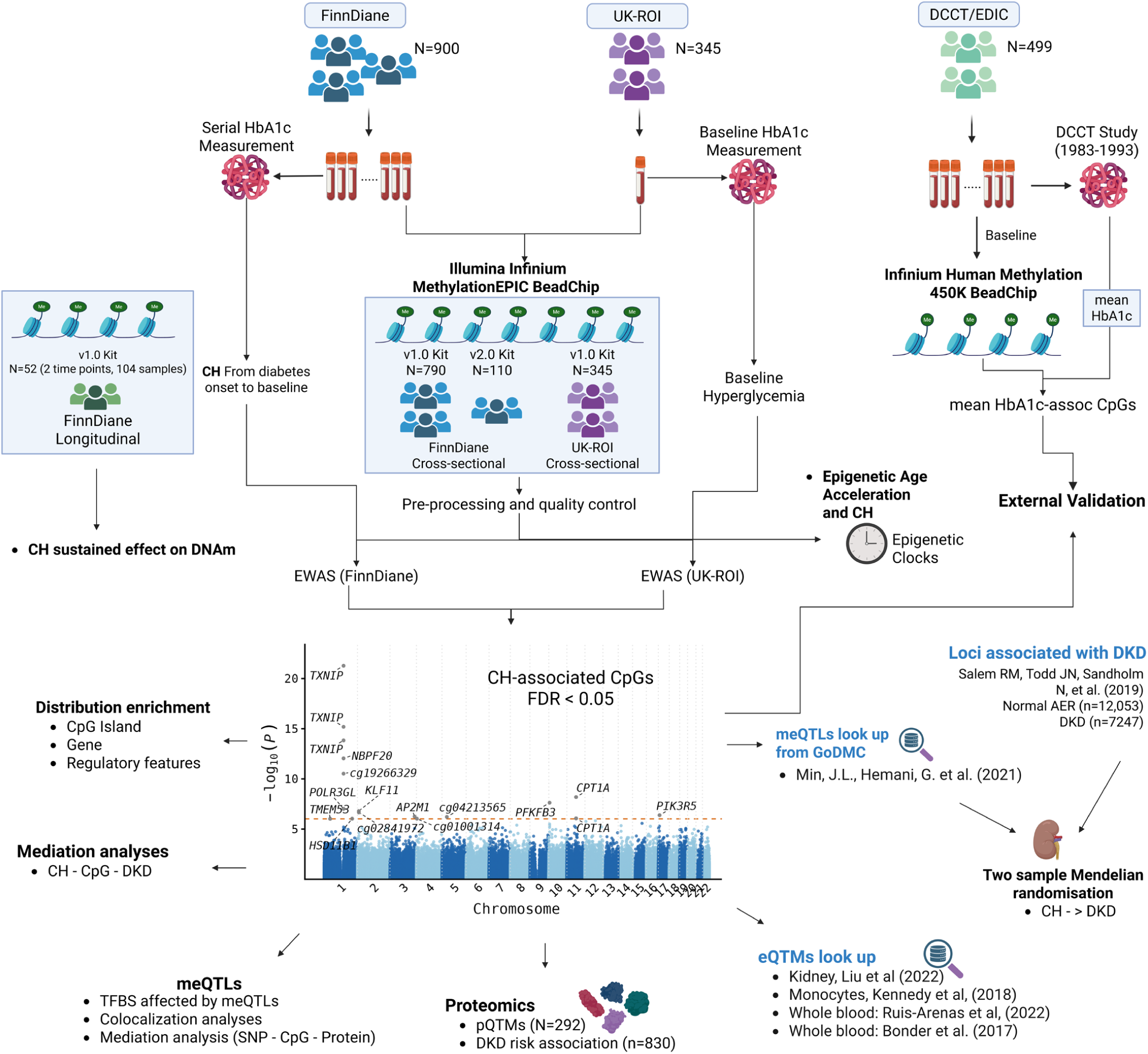
Study design. Created in BioRender. Luo, X. (2026) https://BioRender.com/1y7crgl

## Results

This study included 1245 participants with T1D from the Finnish Diabetic Nephropathy (FinnDiane; n = 900) and United Kingdom-Republic of Ireland (UK-ROI; n = 345) studies (**Fig. 1**). For the FinnDiane cohort, samples were profiled using Infinium MethylationEPIC BeadChips v.1.0 (EPICv1, n = 790) or v.2.0 (EPICv2, n = 110). The participants in EPICv1 group included 62.4% males with a mean age of 42.2 years and diabetes duration of 28.5 years, while EPICv2 had slightly older participants (51.8 years) with longer duration (39.1 years) and all with some level of DKD (**Table 1**). Participants in the UK-ROI cohort were profiled with MethylationEPIC BeadChip v.1.0 and had similar age (41.9) and diabetes duration (26.7) as the FinnDiane EPICv1 participants. CH was defined as cumulative exposure to HbA1c levels above 53 mmol/mol (7%) from diabetes onset to baseline.

**Table 1:**
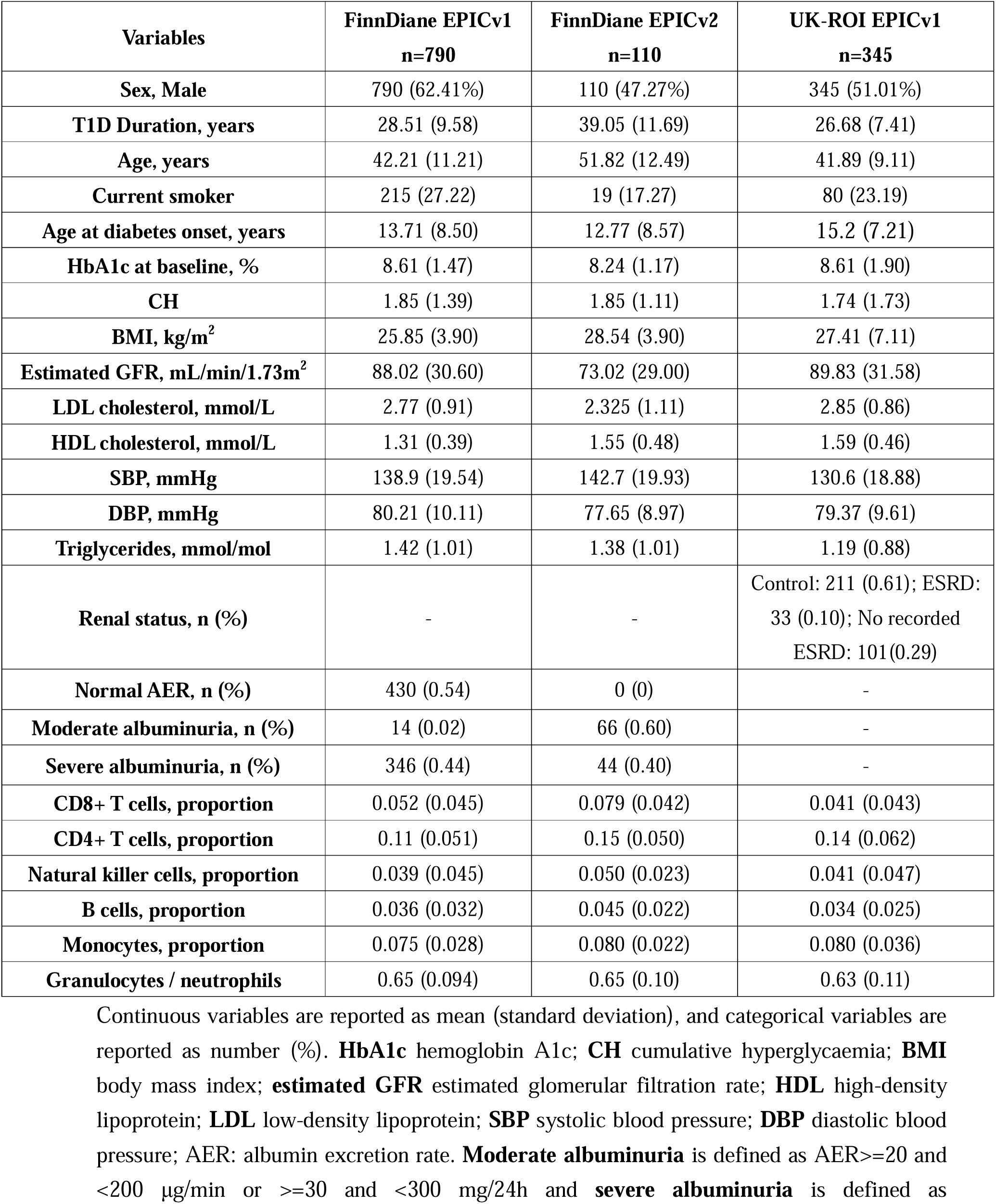
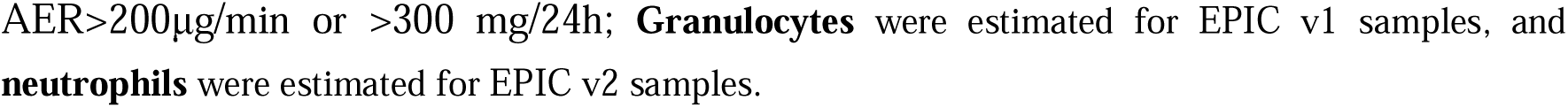
Clinical characteristics of the FinnDiane and UK-ROI study participants.

| Variables | FinnDiane EPICv1<br>n=790 | FinnDiane EPICv2<br>n=110 | UK-ROI EPICv1<br>n=345 |
| --- | --- | --- | --- |
| **Sex, Male** | 790 (62.41%) | 110 (47.27%) | 345 (51.01%) |
| **T1D Duration, years** | 28.51 (9.58) | 39.05 (11.69) | 26.68 (7.41) |
| **Age, years** | 42.21 (11.21) | 51.82 (12.49) | 41.89 (9.11) |
| **Current smoker** | 215 (27.22) | 19 (17.27) | 80 (23.19) |
| **Age at diabetes onset, years** | 13.71 (8.50) | 12.77 (8.57) | 15.2 (7.21) |
| **HbA1c at baseline, %** | 8.61 (1.47) | 8.24 (1.17) | 8.61 (1.90) |
| **CH** | 1.85 (1.39) | 1.85 (1.11) | 1.74 (1.73) |
| **BMI, kg/m<sup>2</sup>** | 25.85 (3.90) | 28.54 (3.90) | 27.41 (7.11) |
| **Estimated GFR, mL/min/1.73m<sup>2</sup>** | 88.02 (30.60) | 73.02 (29.00) | 89.83 (31.58) |
| **LDL cholesterol, mmol/L** | 2.77 (0.91) | 2.325 (1.11) | 2.85 (0.86) |
| **HDL cholesterol, mmol/L** | 1.31 (0.39) | 1.55 (0.48) | 1.59 (0.46) |
| **SBP, mmHg** | 138.9 (19.54) | 142.7 (19.93) | 130.6 (18.88) |
| **DBP, mmHg** | 80.21 (10.11) | 77.65 (8.97) | 79.37 (9.61) |
| **Triglycerides, mmol/mol** | 1.42 (1.01) | 1.38 (1.01) | 1.19 (0.88) |
| **Renal status, n (%)** | - | - | Control: 211 (0.61); ESRD:<br>33 (0.10); No recorded<br>ESRD: 101(0.29) |
| **Normal AER, n (%)** | 430 (0.54) | 0 (0) | - |
| **Moderate albuminuria, n (%)** | 14 (0.02) | 66 (0.60) | - |
| **Severe albuminuria, n (%)** | 346 (0.44) | 44 (0.40) | - |
| **CD8+ T cells, proportion** | 0.052 (0.045) | 0.079 (0.042) | 0.041 (0.043) |
| **CD4+ T cells, proportion** | 0.11 (0.051) | 0.15 (0.050) | 0.14 (0.062) |
| **Natural killer cells, proportion** | 0.039 (0.045) | 0.050 (0.023) | 0.041 (0.047) |
| **B cells, proportion** | 0.036 (0.032) | 0.045 (0.022) | 0.034 (0.025) |
| **Monocytes, proportion** | 0.075 (0.028) | 0.080 (0.022) | 0.080 (0.036) |
| **Granulocytes / neutrophils** | 0.65 (0.094) | 0.65 (0.10) | 0.63 (0.11) |

Continuous variables are reported as mean (standard deviation), and categorical variables are reported as number (%). **HbA1c** hemoglobin A1c; **CH** cumulative hyperglycaemia; **BMI** body mass index; **estimated GFR** estimated glomerular filtration rate; **HDL** high-density lipoprotein; **LDL** low-density lipoprotein; **SBP** systolic blood pressure; **DBP** diastolic blood pressure; AER: albumin excretion rate. **Moderate albuminuria** is defined as AER ≥ 20 and <200 μg/min or ≥ 30 and <300 mg/24h and **severe albuminuria** is defined as
AER>200µg/min or >300 mg/24h; **Granulocytes** were estimated for EPIC v1 samples, and **neutrophils** were estimated for EPIC v2 samples.

### Epigenome-wide identification of blood cell DNA methylation sites (CpGs) associated with CH

After quality control, 761,538 CpGs were included in the EWAS meta-analysis on CH across FinnDiane and the UK-ROI cohorts. Association between DNA methylation and CH was tested using *limma* linear regression model, adjusted for baseline age, sex, smoking status, glomerular filtration rate (eGFR), estimated white blood cell counts (WCCs), and technical variables. We identified 17 CpG sites from 10 distinct chromosomal regions significantly associated with CH (FDR<0.05; **Table 2**, **Fig. 2A**). Furthermore, 115 CpG sites reached a suggestive p-value of < 5×10^-5^ (**Supplementary Table 1**), predominantly (42.5%) located in the gene body (**Fig. 2D**). The quantile-quantile plot showed no evidence of genomic inflation (**Fig. 2B**), with a Bacon-estimated inflation factor of 0.92.

**Figure 2.**
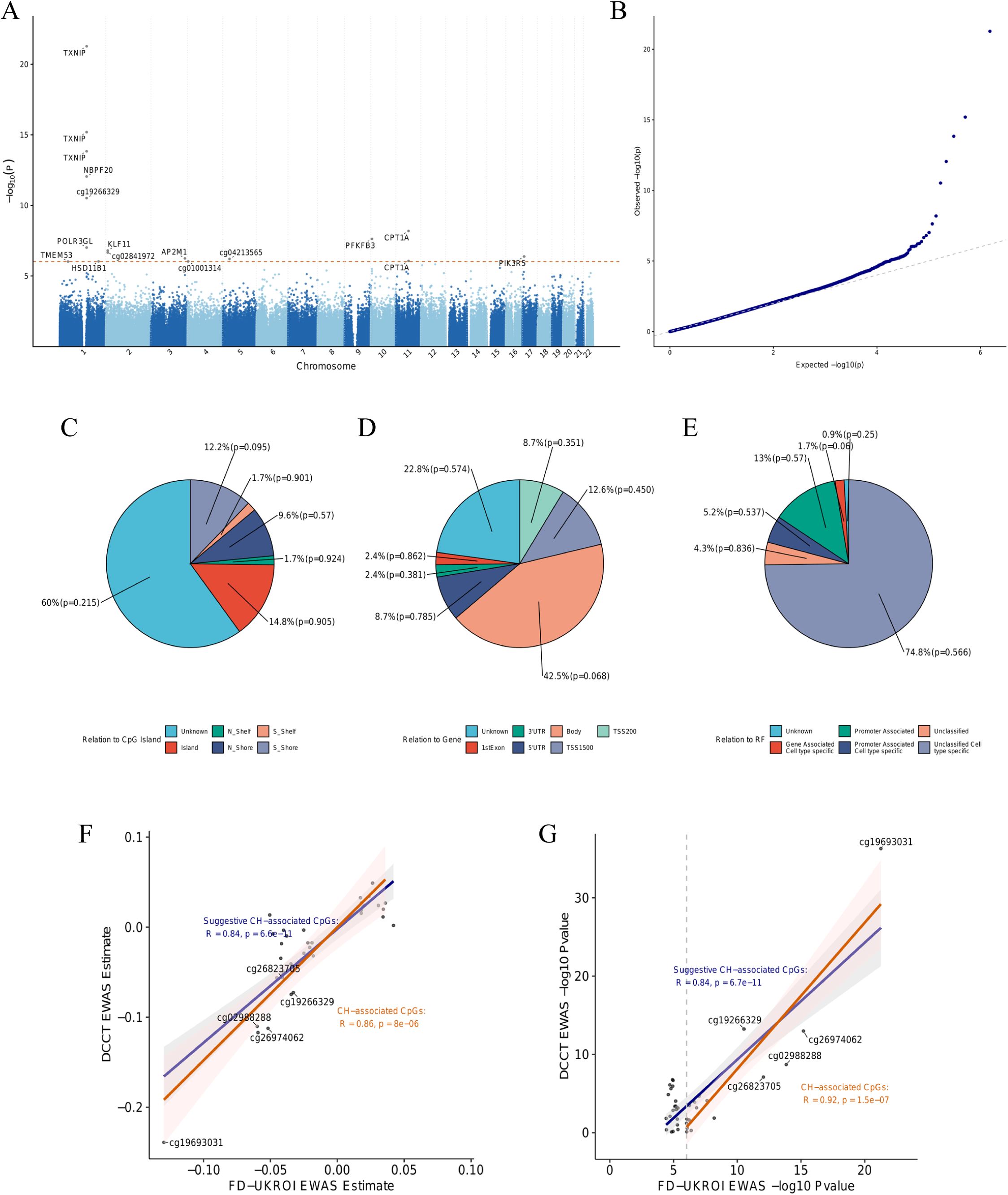
Manhattan **(A)** and qq-plot **(B)** of the epigenome-wide association of cumulative hyperglycaemia (CH) in type 1 diabetes. The location of CH-associated CpGs in relation to CpG island, gene, and regulatory features **(C) (D) (E)**. Spearman correlation of CH-association coefficient **(F)** and-log10 Pvalue **(G)** in the discovery stage meta-EWAS and DCCT/EDIC EWAS for mean HbA1c (N=499).

**Table 2:** CpG sites associated with CH identified by meta-analysis of EWAS.

| CpG | CHR | Location | Gene | Estimate | SE | P-value | FDR | Mean methylation β-values | | |
| --- | --- | --- | --- | --- | --- | --- | --- | --- | --- | --- |
|  |  |  |  |  |  |  |  | FD<br>EPICv1 | FD<br>EPICv2 | UKROI |
| cg19693031 | 1 | 145,441,552 | *TXNIP* | -0.130 | 0.013 | 5.35E-22 | 4.094E-16 | 0.53 | 0.76 | 0.55 |
| cg26974062 | 1 | 145,440,734 | *TXNIP,NBPF20,NBPF10* | -0.052 | 0.006 | 6.43E-16 | 2.459E-10 | 0.052 | 0.11 | 0.05 |
| cg02988288 | 1 | 145,440,445 | *TXNIP;NBPF20,NBPF10* | -0.060 | 0.008 | 1.46E-14 | 3.728E-09 | 0.047 | 0.090 | 0.05 |
| cg26823705 | 1 | 145,435,523 | *NBPF20;NBPF10* | -0.041 | 0.006 | 8.98E-13 | 1.718E-07 | 0.34 | - | 0.37 |
| cg19266329 | 1 | 145,456,128 | chr1q21.1 | -0.033 | 0.005 | 3.02E-11 | 4.617E-06 | 0.46 | 0.61 | 0.49 |
| cg24694018 | 1 | 145,457,621 | *POLR3GL* | -0.025 | 0.005 | 9.70E-08 | 9.276E-03 | 0.51 | 0.61 | 0.53 |
| cg20853880 | 2 | 10184444 | *KLF11* | -0.059 | 0.011 | 1.57E-07 | 1.335E-02 | 0.18 | 0.20 | 0.16 |
| cg02841972 | 2 | 10176151 | chr2p25.1 | -0.025 | 0.005 | 2.20E-07 | 1.683E-02 | 0.40 | 0.54 | 0.41 |
| cg05325763 | 11 | 68607719 | *CPT1A* | -0.042 | 0.007 | 6.51E-09 | 8.298E-04 | 0.052 | 0.10 | 0.05 |
| cg08994060 | 10 | 6214026 | *PFKFB3* | -0.041 | 0.007 | 2.35E-08 | 2.571E-03 | 0.25 | 0.30 | 0.24 |
| cg03334071 | 17 | 8860431 | *PIK3R5* | -0.048 | 0.010 | 4.17E-07 | 2.901E-02 | 0.89 | 0.93 | 0.91 |
| cg17343451 | 3 | 183899704 | *AP2M1* | 0.020 | 0.004 | 5.66E-07 | 3.609E-02 | 0.56 | 0.65 | 0.57 |
| cg04213565 | 5 | 35900039 | *chr5p13.2* | 0.036 | 0.007 | 6.17E-07 | 3.628E-02 | 0.19 | 0.35 | 0.20 |
| cg17058475 | 11 | 68607737 | *CPT1A* | -0.042 | 0.008 | 8.61E-07 | 4.284E-02 | 0.040 | 0.077 | 0.04 |
| cg01001314 | 4 | 3669900 | *4p16.3* | -0.040 | 0.008 | 9.28E-07 | 4.284E-02 | 0.84 | 0.88 | 0.85 |
| cg09366519 | 1 | 209877970 | *HSD11B1* | -0.022 | 0.004 | 9.29E-07 | 4.284E-02 | 0.73 | 0.81 | 0.75 |
| cg18315935 | 1 | 45120295 | *TMEM53* | 0.034 | 0.007 | 9.52E-07 | 4.284E-02 | 0.82 | 0.93 | 0.86 |

**CHR** Chromosome; **Location** Genomic position (hg19); **Gene** annotated gene(s) based on Infinium MethylationEPIC v1.0 B4 Manifest File; **Estimate** estimate from the meta-analysis, representing the change in DNA methylation M value per one-unit increase in CH; **SE** standard error of the effect estimate; **Pvalue** association significance between DNA methylation level and CH; **FDR** P value adjusted for multiple testing using false discovery rate (FDR) correction.

The strongest association signal was observed at cg19693031, located in the 3’UTR of the *TXNIP* gene (β=-0.13, 95% CI: [-0.16,-0.10], p = 5.35×10^-22^; **Table 2**). The same cg19693031 has previously been associated with HbA1c^8^, type 2 diabetes, other glycaemic traits, and the metabolic syndrome^12^. In addition, five other CpGs on the *TXNIP* region (annotated to *NBPF20, NBPF10 and POLR3GL*) were significantly associated with CH (**Table 2**). However, these associations were not significant after further adjustment for cg19693031 methylation level, suggesting their association may not be independent of the primary cg19693031 signal (**Supplementary Table 2**). Outside the *TXNIP* region, 7 of the 17 CH-associated CpGs were novel, i.e., not previously reported for HbA1c, insulin resistance, metabolic syndrome or diabetes.

Seven of the seventeen CH-associated CpGs overlapped with transcription factor (TF) binding sites (**Supplementary Table 3**): For example, cg17058475 (*CPT1A*) and cg19266329 overlapped with a binding motif for PPARG, a transcription factor that regulates target genes involved in insulin sensitivity, inflammatory pathways, lipid metabolism, and adipocyte differentiation^13^. On the *TXNIP* region, cg26823705 (annotated to *NBPF20* and *NBPF10*) overlapped with binding motifs for FOXO3 and FOXO4, members of the Forkhead Box O (FOXO) family, involved in metabolic processes^14^. Many of the 17 CH-associated CpGs were previously reported to be associated with smoking (13 CpGs, 76.4%), metabolic syndrome (6 CpGs, 35.3%), and aging (5 CpGs, 29.4%) in the EWAS Atlas (**Supplementary Table 4**).

Sensitivity analyses incorporating four additional baseline clinical variables (systolic blood pressure (SBP), LDL cholesterol, HDL cholesterol, and BMI) showed only a limited number of CpG sites associated with these clinical variables **(Supplementary Fig. 1A**). The inclusion of any single additional covariates resulted in less than 15% change in CH-associated CpGs effect size (**Supplementary Fig. 1B**).

### Persistence of CH-associated CpGs

We observed no statistically significant differences in DNA methylation levels at the 115 suggestively CH-associated CpGs measured across two different time points, median 7.5 years (range: 3.6-16.4 years) apart, in 104 samples from 52 individuals, indicating that methylation at CH-associated CpGs remain stable over time (**Supplementary Fig. 2A**). At the same time, the CH decreased between visits (**Supplementary Fig. 2B**), suggesting a long-lasting epigenetic imprint of hyperglycaemia, in line with the phenomenon of metabolic memory.

### Cross-lagged analysis for the CH-associated CpG site

To further investigate the temporal relationship between DNA methylation and CH, we performed a cross-lagged panel analysis on the longitudinal data of 52 individuals with EWAS and clinical data at two timepoints with an average 7.5 year difference. This analysis was used to examine whether baseline DNA methylation predicts follow-up CH calculated between the time points, or whether baseline CH predicts DNA methylation at the second time-point. Significant (FDR<0.05) cross-lagged effects were seen from baseline DNA methylation level at cg17058475 (5’UTR *CPT1A*) to follow-up CH (FDR=0.033) and effects from baseline CH to follow-up DNA methylation level at cg05325763 (5’UTR *CPT1A*; FDR=0.040) (**Supplementary Table 5**). Moreover, the autoregressive path (CpG_baseline_-> CpG_follow-up_) of cross-lagged analysis also suggests that methylation levels at each CH-associated CpGs remain stable over time.

### CH and epigenetic aging

Five of the 17 CH-associated CpGs have been previously reported to be associated with aging according to the EWAS Atlas^15^ (**Supplementary Table 4**), implicating an association between CH and aging. To further evaluate this relationship, we calculated intrinsic epigenetic age acceleration (IEAA) using six established epigenetic clocks. CH was significantly associated with accelerated IEAA in three clocks: DunedinPACE, PhenoAge, and GrimAge in the FinnDiane and UK-ROI meta-analysis (p < 0.05, Fig. 3A). These results suggest that persistent hyperglycaemia is associated with accelerated biological aging.

**Figure 3.**
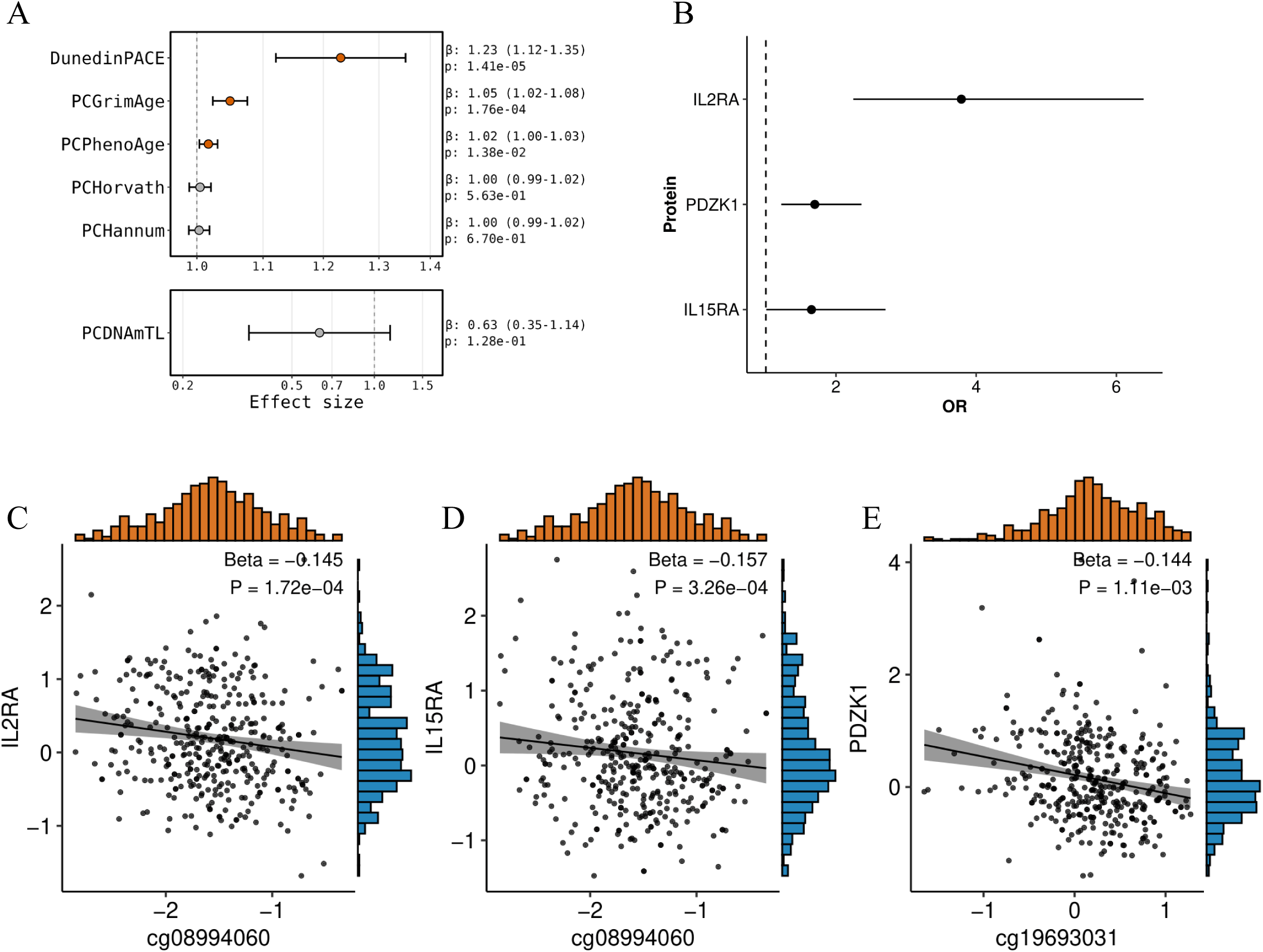
**(A)** Association between cumulative hyperglycaemia and PC adjusted intrinsic epigenetic age acceleration (IEAA) calculated based on DundinPACE, PhenoAge, GrimAge, Horvath, Hannum and DNAmTL epigenetic ageing clocks. **(B)** Forest plot for association between serum protein levels (IL2RA, PDZK1, IL15RA) and DKD. The x-axis shows the Odd Ratio (OR) and 95% confidence interval for DKD per SD change in NPX for each protein. **(C-E)** Scatter plots showing the association between DNA methylation M levels and normalized protein expression levels (NPX) across all 277 samples. **(C)** cg08994060 methylation (*PFKFB3* locus) versus IL2RA expression. **(D)** cg08994060 methylation (*PFKFB3* locus) versus IL15RA expression. **(E)** cg19693031 methylation (*TXNIP* locus) versus PDZK1 expression

### Functional enrichment analysis of the genomic regions at CH-associated CpGs

To study the functional activity of the CH-associated CpGs, we conducted enrichment analysis on regulator regions of 27 major blood cell types from the ChromHMM Study^16^ based on the 115 suggestive CH-associated CpGs (p < 5×10^-5^). We observed prominent enrichment of enhancers and flanking active TSS across 9 major blood cell types (**Supplementary Fig. 3**), indicating widespread active regulatory potential for these CpGs. In contrast, suggestive CH-associated CpGs were significantly depleted in active TSS regions in most cell types, and in repressed polycomb regions in monocytes and hematopoietic stem cells.

Kyoto Encyclopedia of Genes and Genomes (KEGG) and Gene Ontology (GO) pathway enrichment analysis of the genes nearest to the 115 suggestive CH-associated CpGs showed enrichment of multiple pathways related to N-methyl-D-aspartate glutamate receptor activity, growth hormone receptor signaling via JAK-STAT, retinoic acid signaling, resolution of D-loop structures through Holliday junction intermediates, and positive regulation of interleukin-5 production (individual pathway with Bonferroni-corrected p-value <0.05; **Supplementary Fig. 4**).

### Functional association landscape between CH-associated CpGs to genetic variant, gene and protein expression (meQTL, eQTM and pQTM)

We next examined the CH-associated CpGs by integrating genetic variants, gene, and protein expression data. In the FinnDiane, twenty independent *cis*-meQTLs (r^2^<0.05) were identified for five of the 17 CH-associated CpGs, including cg08994060 (*TXNIP*, leading *cis*-meQTL rs7915514), cg20853880 (*KLF11*, leading *cis*-meQTL rs72786623), cg03334071 (*PIK3R5*, *cis*-meQTL rs7218886), cg04213565 (chr5p13.2, *cis*-meQTL rs6451234), and cg18315935 (*TMEM53*, *cis*-meQTL rs2281759) (**Fig. 4A-E**). Nine of the twenty *cis*-meQTL were previously reported by the Genetics of DNA Methylation Consortium (GoDMC)^17^, while 11 were novel. Furthermore, we replicated the robust trans-meQTLs rs35022307 and rs34964576 (intron variants of *SLC2A1*) for cg19693031 (*TXNIP*) previously reported in the general population^11,17,18^ and diabetes cohort^19^. These variants were associated with *SLC2A1-AS1* (*SLC2A1* divergent transcript) expression in the kidney and whole blood in the GTEx (**Extended Data Fig. 1**).

**Figure 4.**
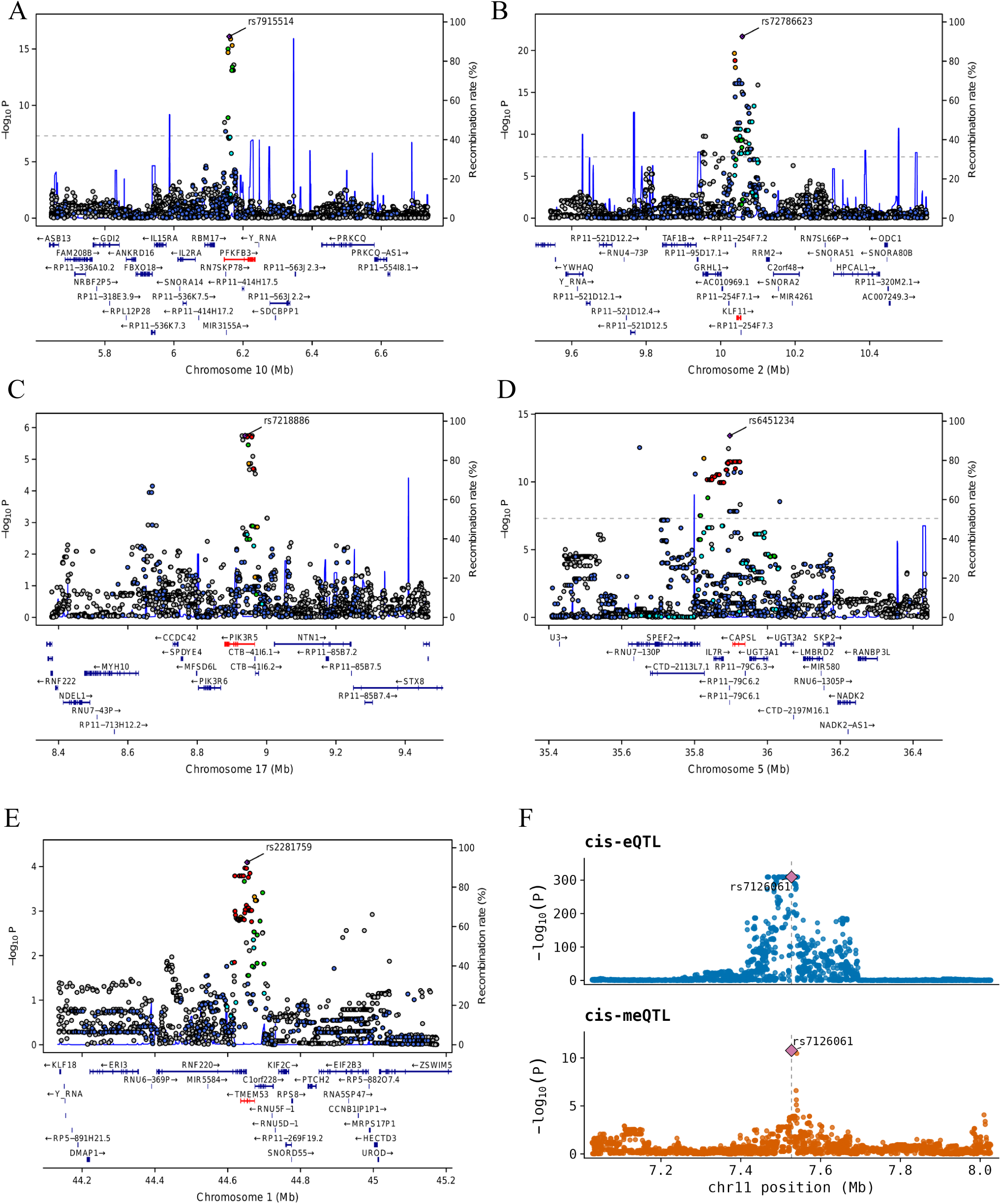
**(A)-(E)** Regional association plots for five representative loci showing SNP associations with CH-associated CpGs, including rs7915514 (lead meQTL for cg08994060, *PFKFB3* locus), rs72786623 (lead meQTL for cg20853880, *KLF11* locus), rs7218886 (lead meQTL for cg03334071, *PIK3R5 locus*), rs6451234 (lead meQTL for cg04213565, *chr5p13.2 locus*), and rs2281759 (lead meQTL for cg18315935, *TMEM53 locus*). X-axis represents genomic position (Mb), left y-axis represent-log10 Pvalue of association. Recombination rates are shown as blue lines and correspond to the right y-axis. The highlighted red gene indicates the nearest gene to lead SNP. **(F)** Regional association plots colocalizing eQTL and meQTL.

We assessed the phenotypic relevance of these meQTLs using the T1D Knowledge Portal for CH-associated meQTLs (**Supplementary Table 7**). The lead meQTL (rs7915514) for cg08994060 (*PFKFB3*) showed associations with T1D, glycaemic traits, and kidney outcomes. For cg20853880 (gene body of *KLF11*), the lead meQTL (rs4669468) was associated with diabetic retinopathy; kidney traits including serum urate, uric acid, and posterior urethral valves; and glycaemic traits (fasting hypothyroidism, acute insulin, and disposition index). rs6451234 (meQTL for cg04213565) was linked to diabetic nephropathy and diabetic retinopathy.

We extended the meQTL analysis to 115 suggestive CH-associated CpGs and identified 120 additional independent *cis*-meQTLs and 9 trans-meQTLs. Among these, 42 meQTLs were previously reported (**Supplementary Table 6**). Furthermore, 78 of the 140 *cis*-meQTLs were strongly disruptive of TF binding motif affinity supporting their functional relevance (**Supplementary Table 8**).

In terms of downstream transcriptional effect, ten eQTM genes for ten out of 17 CH-associated CpGs were identified in kidney^20^, whole blood^21,22^ and monocytes^23^ (**Supplementary Table 9**). *TXNIP* expression was associated with 4 out of 6 lead CpGs at the wider *TXNIP* locus, including cg19693031 (3’UTR of *TXNIP*), cg26974062 (*TXNIP* gene body), cg02988288 (*TXNIP* 5’UTR), and cg19266329 (downstream of *POLR3GL*). Additionally, DNA methylation at cg05325763 and cg17058475 (both 5’UTR of *CPT1A*) were *cis*-eQTMs in the kidney/whole blood for *CPT1A,* a potential therapeutic target for DKD^24^. Furthermore, cg20853880 (gene body *KLF11*) was a *cis*-eQTM for *KLF11* in whole blood and monocytes. Methylation at cg03334071 (*PIK3R5*) was associated with the expression of *NTN1*, an early biomarker of tubular damage, reported to lead to chronic kidney disease (CKD) progression^25^, and its main meQTL rs56255657, was associated with both glycaemic and kidney traits.

To extend these findings to protein level, we performed *cis*-pQTM analysis, i.e. CpG association with protein levels, in 292 FinnDiane participants with DNA methylation and serum proteome data of >5,400 proteins available at the same time point. While TXNIP protein expression was not captured on our proteomics panel, the CH-associated CpG sites on the *TXNIP* locus had cis-pQTM associations with PDZK1 and HJV (**Supplementary Table 10**). cg08994060 at *PFKFB3* locus was associated with IL2RA and IL15RA protein expression. Notably, three of the *cis*-pQTM target proteins, namely IL2RA (*cis*-pQTM of cg08994060, **Fig. 3C**), IL15RA (*cis*-pQTM of cg08994060, **Fig. 3D**), and PDZK1 (*cis*-pQTM of cg19693031 **Fig. 3E**) were significantly associated with DKD (**Fig. 3B**). Serum levels of PDZK1 protein were strongly associated with triglycerides (beta = 0.20, p = 8.40×10^-14^) in the FinnDiane cohort. Furthermore, we identified altogether twenty-seven *cis*-pQTMs associated with seventeen out of 115 suggestive CH-associated CpGs (FDR<0.05; **Supplementary Table 10**).

### Gene expression co-localization analysis

To further investigate potential gene expression patterns affected by the lead CpGs, we conducted colocalisation analysis for altogether 176 eQTL-meQTL pairs based on blood eQTL genes (eQTLGen phase I^26^) for 140 FinnDiane *cis*-meQTLs of the 115 suggestive CH-associated CpGs (**Supplementary Fig. 5**, **Supplementary Table 11**). Colocalisation analysis indicated a potential colocalisation effect between cg21767391 methylation and *PPFIBP2* expression, with a posterior probability of 72.5%. Regional association plots further confirmed the overlap of the association signals across both traits (**Fig. 4F**). Of note, *PPFIBP2* is highly expressed in the kidney collecting duct intercalated cells, papillary tip epithelial cells, vascular smooth muscle cells, and macrophages (**Supplementary Fig. 6**). Additionally, we detected moderate evidence of colocalisation for *RP11-660L16.2* and cg04774822, *CTSW* and cg27305772, and *C19orf52* and cg15224011 with posterior probabilities of 64.9%, 54.1%, and 51.9%, respectively.

Further mediation analysis integrating FinnDiane proteomics data demonstrated that DNA methylation at cg27305772 significantly mediated the effect of rs501630 on CTSW serum protein levels (**Supplementary Table 12**).

### Validation of CH-associated CpGs in an external cohort

We selected 32 CpG sites for validation in the DCCT/EDIC T1D cohort, including all 17 CpG sites significantly associated with CH (FDR<0.05), and 15 suggestive CH-associated CpGs with additional evidence from the downstream analyses (**Supplementary Table 13**). Ten of the 17 CH-associated CpGs, and nine of the 15 suggestive CpGs were replicated (p<1.56×10^-3^, i.e. 0.05/32, Bonferroni correction). Importantly, the effect sizes (R=0.86, p = 8.00×10^-6^, **Fig. 2F**) and significance levels (R=0.92, p = 1.49×10^-7^, **Fig. 2G**) of these 17 CH-associated CpGs, as well as of the 32 tested CpGs, were strongly correlated between our discovery EWAS meta-analysis and DCCT/EDIC cohort.

### Mediation effect of CH-associated CpGs on DKD

To assess whether DNA methylation changes may statistically mediate the association between CH and DKD, we performed a mediation analysis for the 17 CH-associated CpGs (FDR<0.05). Altogether, 11 CpG sites exhibited suggestive mediation effect for the association of CH with DKD (P_ACME_<0.05 and proportion of mediation>5%) (**Fig. 5A**, **Supplementary Table 14**). Notably, methylation at cg19693031 (*TXNIP* 3’UTR), cg02841972 (chr2p25.1, upstream *KLF11*), cg05325763 (*CPT1A* 5’UTR), and cg26823705 (*TXNIP*) were estimated to account for 22.72%, 13.72 %, 12.52%, and 10.16%, respectively, of the association between CH and DKD.

**Figure 5.**
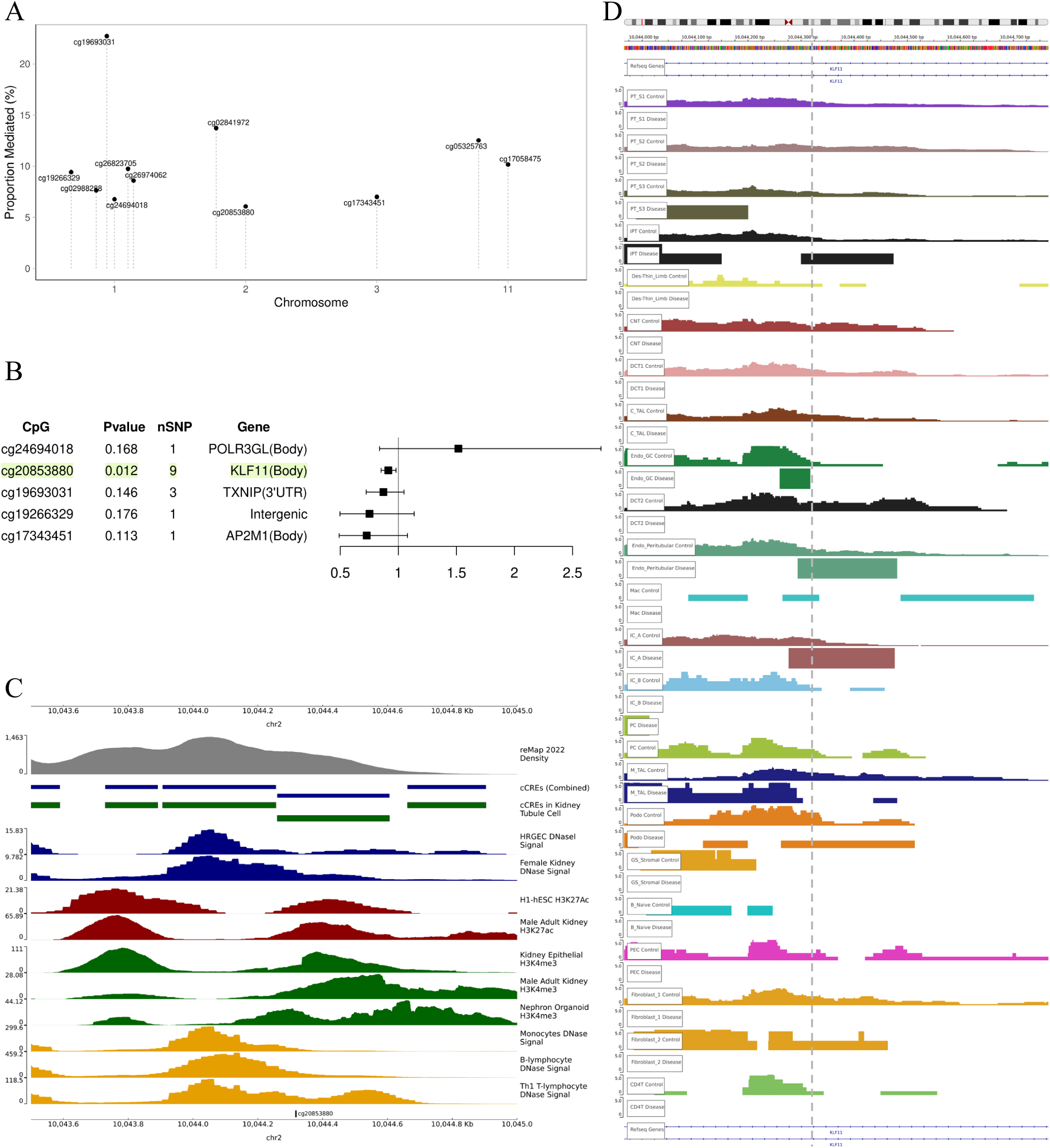
**(A)** Mediation effects of CH-associated CpG methylation linking cumulative hyperglycaemia to DKD. Y-axis: percentage of mediation effect from cumulative hyperglycaemia to DKD explained by methylation at each CpG site. X-axis: genomic position by chromosome. **(B)** Two-sample MR estimates (ORs) from the CH-associated CpG (FDR<0.05) to DKD. The ORs were calculated using IVW method if several mQTL were available for the CpG site or the Wald ratio if only one mQTL was available. **(C)** Epigenomic landscape of cg20853880 in the *KLF11* locus. **(D)** Open chromatin signals in human kidney disease adapted from http://www.susztaklab.com/Human_CKD_snATAC/index.php (chr2:10,043,967-10,044,766, hg38), with cg20853880 (chr2:10044317-10044318, hg38) highlighted. Each track represents chromatin accessibility in different health condition and cell type: endothelial cells of glomerular capillary tuft (Endo_GC), distal convoluted tubule (DCT), proximal tubule (PT) segments 1-3 (PT_S1, S2, S3), injured_PT (iPT), cortical thick ascending loop of Henle (C_TAL), connecting tubule (CNT), thick ascending limb (Des-Thin_Limb), principal cell of collecting duct (PC), endothelial cells (IC), macrophage (Mac), endothelial cells of peritubular vessels (Endo_Peritubular), medullary thick ascending loop of Henle (M_TAL), parietal epithelial cell (PEC), podocyte (Podo), naive B lymphocyte (B_Naive), lymphocytes CD4+ (CD4T), fibroblast (fibroblast), glomerulosclerosis-specific stromal cells (GS_Stromal). Control (n = 36) were defined by eGFR>60 ml min^-1^ 1.73 m^-2^ and fibrosis score of <10%, and disease (n = 45, 20 DKD and 25 hypertensive-attributed CKD) were defined by eGFR< 60 ml min^-1^ 1.73 m^-2^ or kidney fibrosis score of >10%. The full-sized track is shown in Supplementary Figure 9.

### Causal effect of CH-associated CpGs on DKD

To further evaluate the directionality suggested by observational mediation analysis and causal effect of the methylation at the CH-associated CpGs on DKD, we performed two-sample MR analysis using meQTL data from the Genetics of DNA Methylation Consortium (GoDMC; accessed October 2025) and GWAS on DKD in T1D^27^. A total of 23 independent meQTL variants for five CH-associated mediating CpGs were included as instrumental variables for two-sample MR. Methylation at cg20853880 (*KLF11*) showed evidence of a causal effect on DKD (causal OR = 0.91[0.85-0.98], p = 0.012, **Fig. 5B**, **Supplementary Table 15**) under the inverse variance weighted algorithm. Sensitivity analyses did not indicate violations of MR assumptions. Cochran Q statistics showed no evidence of heterogeneity in the MR-Egger (Q = 5.40, p = 0.61) and inverse-variance weighted (IVW) (Q = 5.45, p = 0.71) analyses. Both the MR-PRESSO global test (intercept =-4.0 × 10^-3^, p = 0.83) and the MR-Egger intercept (intercept =-0.004, p = 0.83) were non-significant, indicating no evidence of horizontal directional pleiotropy. Additionally, leave-one-out analysis (**Supplementary Fig. 7A, B**) indicated that the causal effect was stable and not impacted by individual SNPs.

Methylation levels at cg20853880 in FinnDiane (EPICv1, n = 790) were significantly lower in individuals with severe albuminuria compared to those with normal AER (Wilcoxon rank sum test; P = 1.1×10^-4^), and with significant differences across three albuminuria subgroups (Kruskal–Wallis test, P =2.86×10^-5^; **Supplementary Fig. 7C**).

### Regulatory landscape of cg20853880 (*KLF11*) with putative causal role in DKD

Next, we examined whether cg20853880 (*KLF11*) that was causally linked to DKD in MR is located on an active regulatory region in the kidney. Indeed, genomic region around cg20853880, located in the first intron of *KLF11*, is a candidate *cis*-Regulatory Element (cCRE) region with promoter-like signatures identified by SCREEN and active histone marks, including layered H3K4me3, H3K27ac, and H3K4me1 (**Fig. 5C**). The active promoter-proximal regulatory environment is supported by H3K4me3 and H3K27ac signals in blood immune cells, and in kidney and kidney-epithelial cells obtained from ENCODE data (**Supplementary Table 16**). Consistently, single-nucleotide Assay for Transposase-Accessible Chromatin using sequencing (snATAC-Seq) data from Human Kidney Open Chromatin Atlas^20^ showed that cg20853880 is located in an open chromatin region across several cell types in the kidney (**Supplementary Fig. 8**). Comparative analysis of chromatin accessibility between control and CKD samples from the Open Chromatin Atlas of Human Kidney Disease^28^ revealed cell type-specific patterns: Open chromatin signals were detected in controls across almost all cell types **(Fig. 5D, Supplementary Fig. 9**) while absent in kidney disease. On the other hand, endothelial cells of glomerular capillary tuft, intercalated cells type α and injured proximal tubule had stronger peak density in kidney disease compared to controls.

Moreover, the methylation level of cg20853880 was significantly associated with *KLF11* expression in monocytes and whole blood (**Supplementary Table 9**). Thus, together with hypomethylation in DKD and eQTMs, these findings imply that methylation changes at cg20853880 are associated with alterations of chromatin accessibility and potentially influence transcription factor binding and *KLF11* expression. Further, *KLF11* showed significantly lower expression in the early diabetic nephropathy tubulointerstitial samples compared to the samples from healthy living kidney donors (**Supplementary Fig. 7D**), while glomerular samples showed no significant difference (**Supplementary Fig. 7E**). In kidney biopsy samples^29^, *KLF11* expression was significantly reduced in both early and advanced diabetic nephropathy compare with control human kidney samples (**Supplementary Fig. 7F**).

### Single-cell characterization of *KLF11*

Cell type-specific analysis revealed that *KLF11* expression level remained low across most kidney cell populations in the kidney tissue^30^ (**Supplementary Table 17**) and had cell-type specific expression in endothelial cells of glomerular capillary tuft (p = 1.90×10^-12^), medullary thick ascending loop of Henle (p = 4.60×10^-3^), and endothelial cells of peritubular vessels (p = 6.00×10^-7^). Further compared to the control samples, *KLF11* was significantly reduced in endothelial cells of glomerular capillary tuft (p = 4.10×10^-8^) and medullary thick ascending loop of Henle (p = 3.50×10^-2^), suggesting a potential connection between reduced *KLF11* expression in specific cell type.

## Discussion

Here, we report an EWAS on CH in 1,245 participants with T1D from the FinnDiane and UK-ROI cohorts. This study demonstrates the association between blood-derived DNA methylation and CH exposure, providing evidence of sustained epigenetic modifications through longitudinal analysis. To our knowledge, this represents the largest diabetes cohort investigated to date, approximately 2.5-fold larger than previously reported mean-HbA1c EWAS cohorts^8,11,31^. We identified 17 associated CpG sites, with the strongest signal observed at cg19693031, located in the 3’UTR of the *TXNIP* gene. This finding aligns with previous reports where *TXNIP* hypomethylation was strongly associated with glycaemic traits in diabetes^8,31–33^. Furthermore, we identified 16 other CH-associated CpGs, of which seven were novel, and showed significant replication support in the DCCT/EDIC T1D cohort. Integrative analyses combining genomic, epigenetic, and proteomic data further characterized the CpGs. Further, mediation analysis suggested that 11 CpGs mediate the CH effect on DKD, and methylation site cg20853880 in *KLF11* showed significant causal association with DKD in a MR analysis.

The external validation of our EWAS results suggests a consistent association of CH exposure with DNA methylation. The coefficients and significance levels of seventeen CH-associated CpGs showed high correlation between our FinnDiane-UK-ROI meta-analysis and the DCCT/EDIC EWAS. Importantly, 10 of 17 CpGs reached significant replication, including the potential causal site for DKD, cg20853880 (*KLF11*), indicating these signals are reproducible across independent cohorts. While some of our findings, including two CpGs in *CPT1A*, remained non-significant in the replication, the lack of replication may be due to different association models and statistical power rather than false positive findings. While the DCCT/EDIC analysis evaluates the association between DNA methylation and mean HbA1c value, our study focuses on CH exposure, capturing slightly different dimensions of long-term glycaemic traits.

We observed that the methylation levels at the CH-associated CpGs remained stable, despite significant change in CH between the visits in the longitudinal analysis cohort (n=52). This suggests that the epigenetic profile might be shaped primarily by early CH exposure (from diabetes onset to baseline, 28.5 years on average) and remain resistant to short-term (7.5 years on average) fluctuations. This aligns with findings from the DCCT/EDIC study^8^, implying that early intensive glycaemic control is crucial to prevent DKD progression. Surprisingly, our cross-lagged panel model identified a bidirectional effect at the *CPT1A* locus involving two nearby CpG sites cg17058475 (chr11:68607737) and cg05325763 (chr11:68607719). Specifically, baseline methylation at cg17058475 predicted future CH, whereas baseline CH predicted future methylation level of cg05325763. Notably, these specific sites were differentially methylated in DKD individuals with T1D^34^. Furthermore, cg17058475 has been significantly associated with metabolic syndrome^35^ and triglycerides^36^. *CPT1A* encodes the protein carnitine palmitoyltransferase 1A, a key rate-limiting enzyme of fatty acid oxidation, which has been shown to improve albuminuria and glomerular sclerosis^24^. Although these two CpG sites are located only 18bp away from each other, they may serve distinct functions within the *CPT1A* 5’UTR: The cg17058475 overlaps with a V_PPARG_02 motif in kidney tissue, suggesting a potential influence in gene expression regulation, while cg05325763 lies outside this binding site. To our knowledge, this represents the first evidence of a directional effect between CH and DNA methylation in T1D. However, replication of these cross-lagged effect analyses in further cohorts with methylation measured at multiple timepoints is necessary to confirm the finding.

In additional analyses, CH was also associated with accelerated biological aging quantified by DNA methylation clocks. This observation suggests that high glucose exposure enhances biological aging, leaving a persistent influence on the epigenome. Of note, HbA1c was among the clinical biomarkers used to train the third generation clock DunedinPACE^37^, which captures the pace of biological aging. Therefore, the association between CH and DunedinPACE may reflect the extent to which pathologically elevated HbA1c is captured by excess epigenetic ageing beyond chronological age. Hyperglycaemia is linked to oxidative stress, inflammation, and disturbances in proteostasis which act as primary triggers for the aging process^38^. Specifically, hyperglycaemia exposure can lead to increased advanced glycation end-products (AGEs), which are considered as biomarkers of aging and can lead to kidney lesions in diabetes related to kidney aging^39^. Further, accumulated AGEs cause kidney dysfunction by inducing podocyte damage^40,41^.

While CpGs on the *TXNIP* region have been consistently associated with glycaemic traits^31^ and the lead CpG cg19693031 is associated with *TXNIP* mRNA expression, our proteomic analysis identified a novel association between serum PDZK1 and the methylation levels at *TXNIP* CpGs, and with DKD in our data. In the kidney, *PDZK1* is highly expressed in kidney tubular epithelial cells^42^ and its downregulation, mediated by TGF-β1, has been shown to promote kidney fibrosis^43^. In contrast, *PDZK1* overexpression has been reported to inhibit cellular senescence and ameliorate lesions in early-stage diabetic retinopathy^44^. These findings suggest that *PDZK1* dysfunction occurs in both kidney and retinal tissues due to hyperglycaemia. *PDZK1* also found to be associated with triglycerides in the FinnDiane data and the T1D Knowledge Portal, and three SNPs (i33968C > T, i15371G > A, and i19738C > T) in *PDZK1* were identified to associate with lipids and increased risk of the metabolic syndrome^45^.

In our identified *cis*-pQTMs, three (BSG, PLAM, FSTL3) were already previously reported to be associated with the late-stage DKD progression associated CpG, cg00994936, in our recent study^46^. *BSG* is a transmembrane glycoprotein, and its depletion has been shown to impair gluconeogenesis and improve hyperglycaemia in mice^47^. *FSTL3*, a member of the follistatin family, has been proven to be a potential therapeutic target for DKD, suppressing high glucose-induced mesangial cell proliferation and extracellular matrix accumulation by inhibiting the p38 MAPK signaling pathway^48^.

Previous large-scale EWASs have identified DNA methylation signals associated with both hyperglycaemia and diabetes complications^11^ but have identified only a few causal links. For example, the *TXNIP* locus is a well-replicated marker associated with both glycaemic traits^31,49^ and kidney outcomes^50,51^ but it does not seem to be causally linked to DKD^33^. In contrast, the mean HbA1c-associated methylation site cg08309687 within the intron of the non-coding RNA gene *ENSG00000273102* has been shown to have a potential causal effect on eGFR decline^8^. Although *TXNIP* represents the most replicated CH-associated methylation signal, we focused our mechanistic investigation on cg20853880, which showed evidence of a potential causal effect to DKD (**Extended Data Fig. 2**). Interestingly, *KLF11* is known to couple with chromatin pathways and epigenetic regulators, including histone acetyltransferase and HP1^52^. Depending on the *KLF11* transcript isoform, the identified CpG site cg20853880 is located either in the first exon or first intron of *KLF11*. This could be biologically meaningful, because first introns are frequently enriched for active regulatory region chromatin marks^53^ and play critical roles in diseases pathogenesis^54,55^.

*KLF11* is a zinc-finger transcription factor of the Krüppel-like factor (KLF) family. It is a known diabetes-associated gene^56^ and has been implicated in hepatic triglyceride metabolism regulation^57^, lipid^58^, and glucose metabolism, particularly in pancreatic beta cell function^56^ and *KLF11* overexpression inhibits human proinsulin gene expression^59^. However, its function in T1D is rarely described. *KLF*-like factors regulate core physiological processes in the kidney, including maintaining glomerular filtration barrier, tubulointerstitial inflammation, and kidney fibrosis progression^60^. We found that *KLF11* was significantly downregulated in the tubulointerstitium in the early-stage diabetic nephropathy samples compared to the healthy kidney samples, suggesting a loss of *KLF11*-mediated protective function in early-stage DKD. We also observed consistent downregulation of *KLF11* in both glomerular endothelial cells and the medullary thick ascending limb in kidney single cell transcriptome, potentially implying a loss of protective transcriptional regulation in glomerular endothelial cell and medullary thick ascending limb. In addition, the T1D Knowledge Portal suggested strong association between *KLF11* and kidney related biomarkers, including cystatin C and serum urate. Mechanistically, *KLF11* suppresses endothelial activation and inflammation via NF-κB inhibition, reducing IL-6 and endothelin-1 production^61^. In the murine unilateral ureteral obstruction model, *KLF11* deficiency enhanced kidney fibrosis by upregulating the TGF-β/SMAD3 signaling pathway^62^ and pro-inflammatory chemokine pathway. Notably, TGF-β/SMAD3-mediated kidney fibrosis^63^ and NF-κB-regulated^64^ pro-inflammatory pathways (including MCP-1, IL-6, TNF-α, and ICAM-1 gene) are main pathological hallmarks to DKD^65^. Consistent with the previous observation that NF-κB inhibition prevents glomerular inflammation and injury in experimental type 1 diabetes mellitus resembling rats^66^, our findings further support the protective role of *KLF11* against DKD. In clinical context, various *KLF* proteins have been suggested as potential drug targets to prevent kidney injury and slow progression to fibrosis. For instance, statins were suggested to protect against acute kidney injury by increasing *KLF4* expression to suppress inflammation-induced expression of cell adhesion molecules^67^. In addition, statins^68–70^, suberanilohydroxamic acid, tannic acid^71^, and resveratrol^72^ induce endothelial *KLF2* expression to prevent inflammation and are regarded as potential new therapies for kidney fibrosis. This raises the possibility that *KLF11* may be a promising therapeutic target in DKD. Interestingly, dexamethasone can increase KLF11 mRNA and protein levels in cultured neuronal cells^73^, but whether this translates to the kidney requires further experimental validation.

Our study combines two discovery cohorts and represents the largest EWAS to date with longitudinal and integrative multi-omic perspective, linking CH-associated methylation signals to genetic landscape, gene expression, and potential downstream target proteins. However, we recognize some limitations of our study. FinnDiane is an observational cohort with varying follow-up durations for participants. To address this variation, we harmonized the effect of hyperglycaemia over varying diabetes durations. In UK-ROI, CH was estimated with baseline HbA1c as HbA1c values before baseline were not available. This approximation of CH may introduce bias that leads to more conservative association estimates. We therefore used random-effects meta-analysis to account for potential between-cohort heterogeneity arising from differences in CH derivation. We acknowledge that repeated HbA1c measurements collected at regular intervals within a defined time window would allow a more accurate cumulative estimation. Despite this limitation, our external validation based on mean HbA1c identified overlapping signals, suggesting that our analysis captured reproducible cumulative hyperglycaemia-associated epigenetic variation.

DNA methylation was measured in blood samples and we argue that this is a relevant tissue to study effects of CH. However, our study also focuses on DKD, and DNA methylation of whole blood may have limitations in reflecting kidney epigenetic profile and signalling, as the cell composition and the lifespan of cells in the kidneys and blood vary. However, kidney biopsies are invasive, and they are generally undertaken only if individuals with T1D have a clinical suspicion of non-diabetic kidney disease. Encouragingly, previous studies have demonstrated that certain CpG sites identified in blood have been validated in kidney samples. For instance, five of 19 CpG sites associated with eGFR/CKD in blood have also been associated with the degree of renal cortical fibrosis^74^. The *KLF11* expression difference in the diabetic tubulointerstitium supports our interpretation of our key findings from whole blood samples. Although we identified cg20853880 as a promising DKD-causal CpG and observed its association with *KLF11*, functional effects may be mediated by the wider CpG island rather than a single CpG site and further investigation of kidney samples may provide additional insights.

To conclude, our findings demonstrate that hyperglycaemia exposure leads to long-lasting epigenetic modifications and accelerated biological aging. Further, our results align with previous evidence implicating *TXNIP* in hyperglycaemia-related epigenetic changes and highlight a potential causal role for *KLF11* in DKD suggesting a potential basis for drug development.

## Methods

### Study cohorts and cumulative hyperglycaemia calculation

We included 1245 individuals from two T1D cohorts in this study. The majority of the participants were recruited as part of the Finnish Diabetic Nephropathy (FinnDiane) study, an ongoing nationwide multicenter study in Finland comprising nearly 10,000 individuals with T1D. T1D was defined as diagnosed before the age of 40, with insulin treatment started within the first year of diagnosis. During the study visit, blood samples were collected for DNA extraction and laboratory measurements. In this study, we included 900 FinnDiane participants with DNA methylation data for cross-sectional analysis and 52 participants with additional longitudinal DNA methylation measurements at a second time point with a median interval of 7.5 years (range: 3.6–16.4 years). Together with the FinnDiane participants, an additional 345 individuals with T1D from the UK-ROI collection with available baseline HbA1c (%) data were included in the EWAS meta-analysis.

In the FinnDiane cohort, serial blood HbA1c (%) measurements were collected from the study visits and serial laboratory records. On average, participants had 12 HbA1c measurements before the research baseline visit, with a range of 1-113 measurements. The median duration from the first available HbA1c measurement to research baseline visit was 5.1 years (range: 0-33.8 years). The median HbA1c measure frequency was 4 measurements per year (range: 1-16). HbA1c level between the diabetes diagnosis year and the first available measurement record was extrapolated using the time-weighted mean HbA1c of values between the first measurement and the research baseline visit. CH was calculated by multiplying the value of HbA1c exceeding 53 mmol/mol (7%) at each HbA1c measurement time point by the time interval to the next measurement and summing across from diabetes onset to the baseline visit. To account for differences in pre-baseline observation time across participants, the CH was normalized by the observation period. In UK-ROI, longitudinal HbA1c measurements before baseline was not available. Therefore, CH was approximated using the baseline HbA1c exceeding 53 mmol/mol (7%).

### DNA methylation Quality Control and Pre-processing

FinnDiane participants’ blood samples were processed for DNA methylation profiling in three batches. A total of 896 blood samples were analyzed with MethylationEPIC v1.0 BeadChip (Illumina, San Diego, CA, USA) at the Queen’s University of Belfast, UK, with 797 samples in the first batch and 100 samples in the second batch, combined and quality controlled jointly as described previously^46^. Quality control and preprocessing were performed as described in the previous study^34^. In brief, 761,538 out of 866,895 probes remained after removing SNV-enriched probes, cross-reactive probes, no-context probes, and probes on sex chromosomes. After excluding individuals with missing HbA1c records before baseline and unknown smoking status, 790 participants remained (**Supplementary Table 18**). An additional 152 blood-derived DNA samples were analyzed with MethylationEPIC v2.0 BeadChip (Illumina, San Diego, CA, USA) at the Austrian Institute of Technology (Vienna, Austria). Raw.idat file from the Illumina Infinium MethylationEPIC v2.0 array were processed using R package *’SeSAMe’* (version 1.24.0). The preprocessing workflow and the number of probes retained at each step were described in **Supplementary Table 18.** A total of 839,334 probes remained after preprocessing. After excluding participants included in the MethylationEPIC v1.0 dataset, 110 additional participants were retained.

To address technical variability, principal components (PCs) calculated from 225 non-negative control probed red (Cy5) and green (Cy3) signal intensities were applied, with PCs 1-5 accounting for >95% of the variability. Intrapersonal mean methylation (mean M) was calculated from 86,980 invariable CpGs (beta-value range <0.05 in >50% of technical batches) out of 114,204 known invariable CpGs^75^.

UK-ROI samples were analyzed with MethylationEPIC v1.0 BeadChip (Illumina, San Diego, CA, USA), following the same processing protocol as applied to the FinnDiane MethylationEPIC v1.0 cohort. 770,786 probes remained after removing SNV-enriched probes, cross-reactive probes, no-context probes, and probes on sex chromosomes (**Supplementary Table 18**).

White blood cell counts (WCC), including granulocytes, monocytes, B-cells, CD4+, CD8+, and NK cells for MethylationEPIC v1.0 in both FinnDiane and UK-ROI cohorts, were estimated using the Houseman^76^ method from raw.idat files with *’minfi’* R-package’s estimateCellCounts function. In MethylationEPIC v2.0 FinnDiane samples, neutrophils, monocytes, B-cell, CD4+, CD8+, and NK cells were estimated using FlowSorted.Blood.EPIC package.

### Meta-analysis of EWAS

Multiple linear regression models were used to identify CH-associated CpGs, with normalized methylation level (M values) at each CpG site as the dependent variable and CH as the independent variable. EWAS was performed separately in FinnDiane MethylationEPIC v1.0 (N=790), FinnDiane MethylationEPIC v2.0 (N=110), and UK-ROI (N=345) cohorts. In the FinnDiane cohort, covariates included age, sex, WCCs, smoking status, and baseline glomerular filtration rate (eGFR) calculated using the using Inker et al. (CKD-EPI 2021)^77^ formula. Baseline eGFR was included as a covariate to account for kidney status differences across the cohorts and reduce confounding from DKD-related methylation changes. In addition, technical PCs 1-5, mean methylation M value from invariable sites were adjusted to account for technical variability in FinnDiane MethylationEPIC v1.0 subset. In the UK-ROI cohort, covariates included age, sex, WCCs, smoking status, technical PCs 1-5, and mean methylation M value from invariable sites.

Additionally, we conducted a two-step sensitivity test in FinnDiane MethylationEPIC v1.0 for the final covariate selection. We included BMI, SBP, LDL cholesterol, and HDL cholesterol measured at the time of blood sampling in the model to assess their association with DNA methylation at all CpG sites. Each of these four variables was individually added to the model, to estimate their significance and coefficient changes of the suggestive CH-associated CpGs.

The mLiftOver^77^ function from the R package *’SeSAMe’* (version 1.24.0) was used to harmonize probes from MethylationEPIC v2.0 to the MethylationEPIC v1.0 platform to enable cross-platform meta-analysis of the EWAS in MethylationEPIC v2.0 samples. Inverse-variance weighted fixed-effects meta-analysis of FinnDiane sub-cohort summary statistics was performed using METAL (version 2020-05-05), and subsequently meta-analyzed with the UK-ROI cohort with the random-effect model using the R package’*metafor*’ (version 4.8-0). CpG - CH associations with FDR<0.05, corresponding to p-value of 9.5×10^-07^, were considered statistically significant. Infinium MethylationEPIC v1.0 B4 Manifest File was used to annotate the CpGs. We estimated the Bayesian inflation factor using the R package *bacon* (version: 1.34.0). CH-associated CpGs were further explored in the NGDC EWAS Atlas (https://ngdc.cncb.ac.cn/ewas/atlas) to obtain associations with other phenotypes and non-cancer diseases.

### Enrichment analysis of CH-associated CpGs

A hypergeometric test was applied to test enrichment regarding the location of CH-associated CpGs compared to non-CH-associated CpGs. Functional enrichment analysis was performed using ClueGO (v2.5.10) and CluePedia (v1.5.10). We used Cytoscape plugins to assess enrichment of CH-associated CpGs in KEGG pathways and GO terms, including biological process, cellular component, and molecular function. Pathways and terms with a p-value < 0.05 were considered significant.

### Sustained effect of CH

To assess the persistence of CH-associated methylation changes over time, we examined 52 FinnDiane participants who had DNA methylation measurements at two time points with a median interval of 7.5 years (range: 3.6-16.4 years). DNA methylation M values were regressed on sex, age, PCS 1-5, WCC, mean M, and residuals were used in the mediation analysis. Paired sample t-tests were applied to assess the difference in methylation level over time. We evaluated the CH at both time points, normalized by the year unit. CH_t1_ was CH exposure normalized by the time from diabetes onset to the first DNA methylation measurement, and CH_t2_ was cumulative hyperglycaemia exposure normalized by the time from the first to the second measurement. Changes in CH between time points were evaluated with the Mann-Whitney U test.

### Longitudinal association between DNA methylation and Cumulative Hyperglycaemia

A cross-lagged panel model was applied to examine the bidirectional association between DNA methylation and CH in 52 FinnDiane participants with two time point DNA methylation data available. In cross-lagged panel analysis, the coefficient and significance of ρ1 indicate whether the baseline CH predicts the DNA methylation level of CpG sites at the follow-up time point, while ρ2 represents the prediction in the opposite cross-lagged direction.

### Epigenetic clock generation

Epigenetic clocks were generated as previously described^79^. Epigenetic age (EA) and epigenetic age acceleration (EAA) were calculated for FinnDiane and UK-ROI participants using six representative epigenetic clocks spanning three generations: chronological clocks Horvath, and Hannum; biological ageing clocks PhenoAge, GrimAge, and DNAmTL; and age acceleration clock DunedinPACE. The mLiftOver function from the R package’SeSAMe’ (version 1.24.0) was used to harmonize probes from MethylationEPIC v2.0 to the MethylationEPIC v1.0 platform for epigenetic age estimation. Principal-component clock versions from Higgins-Chen et al^80^. were used. Intrinsic EAA (IEAA) was derived as residuals from EAA regressed on age and WCCs for all clocks except for DunedinPACE, which was adjusted only for WCC. Associations between CH and IEAA were assessed adjusting for sex, and additionally for age in DunedinPACE models. Age was added as a covariate in DunedinPACE models. Inverse-variance weighted fixed-effect meta-analysis was performed between cohorts using METAL (version 2020-05-05).

### Regulatory function characterization of CH-associated CpGs

We performed an enrichment analysis of CH-associated CpGs regarding the transcription regulation status. CpGs were annotated using epigenetic states inferred from ChromHMM based on histone modification profiles from the NIH Roadmap Epigenomics. Fifteen chromatin states defined by ChromHMM model included activate TSS (TssA), Flanking Active TSS (TssAFlnk), Transcription at gene 5’ and 3’ (TxFlnk), Strong transcription (Tx), Weak transcription (TxWk), Genic enhancers (EnhG), Enhancers (Enh), ZNF genes & repeats (ZNF/Rpts), Heterochromatin (Het), Bivalent/Poised TSS (TssBiv), Flanking Bivalent TSS/Enh (BivFlnk), Bivalent Enhancer (EnhBig), Repressed PolyComb (ReprPC), Weak Repressed PolyComb (ReprPCWk), and Quiescent/Low (Quies). We focused on 27 blood-related cells (E062, E034, E045, E033, E044, E043, E039, E041, E042, E040, E037, E048, E038, E047, E029, E031, E035, E051, E050, E036, E032, E046, E030, E115, E116, E123, E124), as described in **Supplementary Table 19**. Fisher’s exact test was used to assess the enrichment significance compared to non-CH associated CpG sites.

### Identification meQTL of CH-associated CpGs in the FinnDiane cohort

We investigated whether genetic variants influence DNA methylation at CH-associated CpG sites. The meQTLs were identified from a subset of 758 FinnDiane study participants who had both genotype and DNA methylation data available using the R package *’MatrixEQTL’* (version 2.3). In total, 6,015,118 quality-controlled genotyped or imputed SNVs^81^ with a minor allele frequency (MAF) ≥ 0.05 and imputation info ≥0.80 were included in the analysis. Linear regression model was applied to identify *cis*-(±1 Mb) and *trans*-meQTLs, and was adjusted for age, sex, diabetes duration, WCCs, and PCs 1-5.

For each CH-associated CpG, LD clumping was applied as described in the section “causal effect identification” to identify independent meQTLs (r^2^ < 0.05). Thereafter, both *cis*-and *trans*-meQTLs were queried from the previously reported meQTLs from GoDMC^17^, UK cohorts^18^, and Human Whole Blood mQTL Atlas (CRIC)^11^ to identify the novel meQTLs.

### meQTLs overlap with transcription factor binding motifs

R package *‘motifbreakR’* (version 2.20.0) was used to evaluate the potential consequences of meQTL genetic variants on nearby transcription factor binding motifs. *MotifbreakR* uses position weight matrix to score the differences of TF binding between the reference and alternative alleles for every possible window that includes the variants. The information content (ic) method was selected to estimate the impact of 141 FinnDiane meQTLs on 691 TF binding sites from the JASPAR 2022 database. All meQTLs with an absolute score difference≥1.5 were considered to altering TF binding.

### TF analysis and expression quantitative trait methylation (eQTM)

We used eFORGE-TF (https://eforge-tf.altiusinstitute.org/) to identify TF motifs that overlap with CH-associated CpG sites. We studied the association between CH-associated CpGs and gene expression using published eQTM datasets from whole blood^21,22^, monocytes^23^, and kidney tissue^20^.

### Protein quantitative trait methylation (pQTM)

Serum protein levels of 5,420 proteins were measured by the OLINK® Explore HT assay at the SciLife Lab, Uppsala, Sweden. The samples were randomised on plates based on sex, age, baseline albuminuria status, kidney disease progression, storage time and thaw cycle number. The protein expression data (NPX, Normalized Protein expression; Log2 scale) were corrected for the plate control and intensity-normalized by the lab. Three samples were excluded due to assay failures, and seven outlier samples were excluded as the overall sample median or the sample interquartile range exceeded 3.5 standard deviations of the overall median or interquartile range.

A subset of 292 participants had both proteomics and DNA methylation data measured at the same time point. We conducted pQTM analysis in these participants to identify proteins associated with CH-associated CpGs. DNA methylation levels were residualized using a linear model to account for technical variation and cell composition, including granulocytes, B cells, CD4 T cells, CD8 T cells, monocytes, NK cells, the first five principal components (PC1–PC5), and mean methylation (mean M). Normalized protein expression value (NPX) was used as the dependent variable, residualized methylation value as independent variable, and age, sex, number of serum thawing, and serum storage time as covariates. Proteins were annotated to genes, and protein expression associated with DNA methylation within ±1 Mb was considered a *cis*-pQTM, while those with larger distance between methylation and gene related to the protein were defined as *trans*-pQTM.

### Colocalisation analysis

We investigated whether the same genetic variants influence both gene expression and DNA methylation using colocalisation analysis. First, we obtained genes associated with variants from the eQTLGene phase I data. Next, eQTLs were intersected with the FinnDiane meQTLs to define the CpG-Gene pairs (**Supplementary Fig. 5**).

Colocalisation analysis was performed using the R package *’coloc’* (version 5.2.3) on 68 CpG-Gene pairs. Summary statistics for each trait were extracted for all variants within ±500kb of the sentinel SNP from the eQTL and meQTL datasets. Prior probabilities were defined as SNP associated with gene expressions (p1), methylation (p2), or both (p12). Variants associated with both DNA methylation and gene expression were visualized using R package *’locuscomparer’* (version 1.0.0).

### Mediation analysis of DNA methylation linking meQTL to protein level

Mediation analysis was performed using the R package *’mediation’* (version 4.5.1) on the CpG-Gene pairs examined in colocalisation analysis to investigate which shared genetic variants influence protein expression through DNA methylation (SNP-Meth-Protein, based on 277 FinnDiane samples with the three omics layers available). The protein encoded by the corresponding gene was used to extend the mediation role from the transcriptional level to the protein level. In mediation analysis, the genetic variant was treated as the exposure, methylation as the mediator, and protein expression as the outcome. DNA methylation residuals generated a previous section describing sustained effect of CH estimation were used in the mediation analysis.

In SNP-Meth-Protein analysis, we estimated the protein expression (NPX) on mediator effect through the linear model:

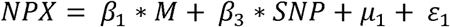

where M is the residual of methylation level at each CpG site,β_1_is the mediator effect on protein expression,β_3_ is the effect of SNP on protein expression,μ_1_is the intercept, and ∈_1_ is the residual error.

The mediator formula

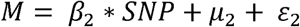

Where β_2_is the effect of genetic variant on DNA methylation, μ_2_is the intercept, and ∈_2_ is the residual error.

Significant mediation effect was considered present when the p-value of ACME < 0.05 and p-value of total effect < 0.05. The proportion of mediation was calculated as following formula:

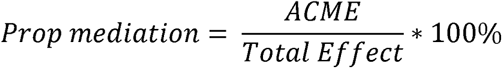

### Replication of CH-associated CpGs in external cohorts

We sought replication of 32 significant or selected suggestive CH-associated CpGs that had significant findings in the downstream analyses (**Supplementary Table 13**) in the Diabetes Control and Complications Trial (DCCT) cohort. The DCCT/EDIC study (n = 499) examined the association between DNA methylation (Infinium HumanMethylation450 BeadChip) and mean HbA1c^8^.

### Mediation analysis to link CH to DKD

The mediation role of CH-associated CpGs in mediating effects of CH to DKD (i.e., CH – CpG - DKD) was studied using the bootstrapping method in the R package *’mediation’* (version 4.5.1). DKD status was defined by albumin excretion rate (AER), as described previously^34^. In brief, moderately increased albuminuria was defined as an AER ≥20 and <200 μg/min or ≥30 and <300 mg/24h and severely increased albuminuria as an AER >200 μg/min or >300 mg/24h in two out of three timed overnight or 24h urine collections. In mediation analysis, DKD cases were defined as those with severely increased albuminuria, and controls as those with normal AER. The CH was treated as the exposure, methylation as the mediator, and binary DKD status as the outcome. Residualized DNA methylation values generated as previously described in the sustained effect of CH estimation were used.

First, we applied linear regression to calculate the mediating effects, and re-estimate the exposure-outcome association adjusting for mediator in the following regression:

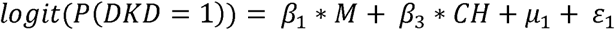

where M is the residualized methylation at each CpG site,β_1_ is the mediator effect on DKD, β_3_ is the effect of CH on DKD,μ_1_is the intercept, and ∈_1_ is the residual error.

The exposure on mediator effect was estimated through regression:

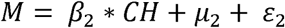

Where β_2_ is the effect of CH on methylation level,μ_2_ is the intercept, and ∈_2_ is the residual error. The indirect effect (average causal mediation effect, ACME) was calculated as β_2_ × β_1_, and the total effect as β_2_×β_1_ +β_3_

CpGs with an ACME p-value<0.05 were considered as significant mediators. For them, the proportion of mediation from CH to DKD by DNA methylation was calculated using the following formula:

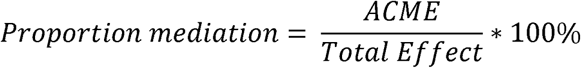

### Causal effect identification

Two-sample MR was conducted to investigate the potential causal effect of DNA methylation at CH-associated CpG sites on the risk of DKD. The study adhered to the STORE-MR guideline based on three core assumptions. Known meQTLs identified in whole blood by the GoDMC were used as instrumental variables. For each CH-associated mediator CpG, *cis*-meQTLs were LD clumped (r^2^<0.05) using linkage disequilibrium estimates from the Finnish population of the 1000 Genomes Project (GRCh37), assessed from ‘*LDlinkR’* R packages (version 1.4.0). Within each LD block, the variant with the strongest association with CpGs was retained. To avoid weak-instrument bias, we excluded meQTLs with *F*-statistics < 10. For outcome data, we retrieved genetic variants associated with DKD (including moderate albuminuria, severe albuminuria, and end stage kidney disease) from the largest GWAS study on DKD in T1D by Salem, et al^27^. Causal effects for each CH-associated mediator CpG were estimated using the R package *’TwoSampleMR’* (version 0.6.14). The Wald ratio method or the IVW method was applied for CpGs with single and multiple instrumental variables, respectively. Thereafter, heterogeneity, pleiotropy, and leave-one-out sensitivity analysis were then performed to confirm the causality (**Supplementary Fig. 10**).

### Epigenomic annotation and regulatory element analysis

To characterize the regulatory role of the *KLF11* cg20853880 CpG site implicated as causal for DKD in the MR analysis, we examined the overlap of CpG genomic location with epigenomic signatures in both general and kidney-specific tissues from SCREEN (Search Candidate Regulatory Elements by ENCODE) and visualized them using pyGenomeTracks (https://github.com/deeptools/pyGenomeTracks). We integrated multiple epigenomic layers described in **Supplementary Table 16**.

### Kidney single-cell level chromatin state and gene expression

We assessed chromatin accessibility at the genomic location of the *KLF11* cg20853880 CpG causally linked to DKD using Human Kidney Open Chromatin Atlas (https://susztaklab.com/Human_snATAC)^20^. Comparative chromatin accessibility difference between DKD samples and healthy controls was obtained from the Open Chromatin Atlas of Human Kidney Disease (http://www.susztaklab.com/Human_CKD_snATAC/index.php)^28^. Further, we evaluated the gene expression of *KLF11*, the gene closest to a CpG causally linked to DKD, in healthy control and DKD samples, from Human Kidney Single Cell Transcriptome (http://www.susztaklab.com/hk_genemap/scRNA)^30^.

### RNA-Seq

To better understand the expression status of *KLF11* in diabetes, we obtained publicly available transcriptome dataset with glomeruli and tubulointerstitial tissue by *Levin et al*^82^. from individuals with biopsy-verified early-stage diabetes (N_T1D_=5 and N_T12D_=14) and living donors (n=20). The raw count data was obtained from KaroKidney. To remove low-abundance transcripts, we only kept genes with read count larger than 10 in at least two samples. Normalization was performed using R package *‘DESeq2’* (version 1.46.0). The “median of ratios” normalization function^83^ calculates read counts relative to the geometric mean per gene. Differential expression analysis was conducted with *DESeq2,* which models gene-level count data with negative binomial distribution. To further evaluate the KLF11 expression level in DKD progression, we then compare the difference in control, early-stage DKD, and advanced-stage DKD in 36 human kidney biopsy samples^84^. To remove low-abundance transcripts, we only kept genes with a normalized count larger than 10 in at least two samples. Overall difference in *KLF11* expression was tested using a likelihood ratio test, and pairwise comparisons between groups were performed using the Wald test.

## Data Availability

Publicly available datasets used in this study include:

EWAS Atlas: https://ngdc.cncb.ac.cn/ewas/atlas

Human Kidney snATAC-Seq: http://www.susztaklab.com/Human_snATAC/index.php

Human Kidney Disease snATAC-Seq: http://www.susztaklab.com/Human_CKD_snATAC/index.php

scRNA-Seq: http://www.susztaklab.com/hk_genemap/scRNA

SCREEN: https://screen.encodeproject.org/

GoDMC meQTL: http://www.godmc.org.uk/

GWAS study of DKD summary statistic from Salem RM, et al.:

https://personal.broadinstitute.org/ryank/JDRF_DNCRI_allvcntrl_min_meta_June2019_rsID. txt.zip

TF: https://eforge-tf.altiusinstitute.org/

eQTL: https://eqtlgen.github.io//eqtlgen-web-site/index.html

GTEx portal: https://gtexportal.org/home/

Type 1 diabetes knowledge portal: https://t1d.hugeamp.org/

Human kidney RNA sequencing data: https://karokidney.org/rna-seq-dn/ and https://www.ncbi.nlm.nih.gov/geo/query/acc.cgi?acc=GSE142025

The human protein atlas: https://www.proteinatlas.org/

Epigenetic states inferred from ChromHMM and samples metadata:

https://egg2.wustl.edu/roadmap/data/byFileType/chromhmmSegmentations/ChmmModels/co reMarks/jointModel/final/all.mnemonics.bedFiles.tgz

https://docs.google.com/spreadsheets/d/1yikGx4MsO9Ei36b64yOy9Vb6oPC5IBGlFbYEt-N 6gOM/edit?gid=15#gid=15

Data collected for the DCCT/EDIC study through June 30th, 2022, are available to the public through the NIDDK Central Repository (https://repository.niddk.nih.gov/studies/edic/). Data collected in the current cycle (July 2022-June 2027) will be available to the public within two years after the end of the funding cycle.

Individual-level data for the study participants (participant-level genome-wide CpG methylation and clinical data for participants in FinnDiane and UK-ROI) are not publicly available due to ethical and legal reasons and the informed consent provided by the FinnDiane participants at the time of data collection.

The FinnDiane study protocol was approved by the ethics committee of the Helsinki and Uusimaa Hospital District (HUS) (491/E5/2006, 238/13/03/00/2015, and HUS-3313-2018, July 3rd, 2019). The ethical approval reference number for the UK-ROI samples are MREC 175/23 (RO321), ORECNI 08/NIR03/79, 12/NI/0003, and 12/NI/0178. The study was performed in accordance with the Declaration of Helsinki.

## Code Availability

No custom code or algorithm was developed for the analyses performed in this study. Previously designed, publicly available pipelines and code were used (as described in the Methods), including:

minfi (https://bioconductor.org/packages/release/bioc/html/minfi.html)

SeSAMe (https://github.com/zwdzwd/sesame),

METAL (https://github.com/statgen/METAL),

metafor (https://cran.r-project.org/web/packages/metafor/index.html),

bacon (https://www.bioconductor.org/packages/release/bioc/html/bacon.html),

PC-Clocks (https://www.bioconductor.org/packages/release/bioc/html/bacon.html), MatrixEQTL (https://github.com/andreyshabalin/matrixeqtl),

LDlinkR (https://cran.r-project.org/web/packages/LDlinkR/vignettes/LDlinkR.html),

motifbreakR (https://bioconductor.org/packages/release/bioc/html/motifbreakR.html),

locuszoomr (https://cran.r-project.org/web/packages/locuszoomr/index.html),

mediation (https://cran.r-project.org/web/packages/mediation/index.html),

coloc (https://chr1swallace.github.io/coloc/articles/vignette.html),

TwoSampleMR (https://mrcieu.github.io/TwoSampleMR/).

## Supporting information

STROBE check list

## Acknowledgements

We thank all individuals who participated in this study. The skilled technical assistance of Anna Sandelin, Anu Dufva, Kirsi Uljala, Heli Krigsnman, and Hanna Olanne (Folkhälsan Research Center and University of Helsinki, Finland) is gratefully acknowledged. We acknowledge all physicians and nurses at each FinnDiane study center participating in the collection of the data (**Supplementary Table 20**). We would like to acknowledge the GENIE consortium collaborators for their contribution in study conceptualization (**Supplementary Table 21**). The authors wish to thank the Finnish Computing Competence Infrastructure (FCCI) for supporting this project with computational and data storage resources for the FinnDiane data; and the ELIXIR Finland node hosted at CSC – IT Center for Science for ICT resources that enabled computational analysis. The gene expression data of *SLC2A1-AS1* and *AKR1C1* in whole blood described in this manuscript were obtained from the GTEx Portal on 01/2026. The Genotype-Tissue Expression (GTEx) Project was supported by the Common Fund of the Office of the Director of the National Institutes of Health, and by NCI, NHGRI, NHLBI, NIDA, NIMH, and NINDS.

## Author contributions statement

XL contributed to the study design, analyzed the FinnDiane data, performed the downstream analyses, and wrote the first draft of the manuscript. AS contributed to the study design, preprocessed FinnDiane EWAS data, contributed to interpretation, and commented on the manuscript. CH preprocessed the UK-ROI EWAS data, performed the EWAS, designed the epigenetic age acceleration analyses and performed those for the UK-ROI cohort. LJS contributed to the generation and quality control of EPICv.1 EWAS data for FinnDiane and UK-ROI. EHD preprocessed the FinnDiane EPICv2 data and contributed to the FinnDiane EPICv1 data quality control. SM contributed to sample selection and processing of the FinnDiane proteomics data. ZC and RN contributed to the validation of results in the DCCT/EDIC EWAS. SP and KS investigated the ATAC-seq regulatory landscape of the DKD-causal CpG sites. AP and HJ contributed to the generation of the FinnDiane EPICv2 data. JNH and JCF contributed to conceptualization of the study and obtaining funding for the GENIE Consortium. GM and APM contributed to the data acquisition and interpretation of the study results and the analysis of the UK-ROI cohort data. P-HG contributed to conceptualization of the study, data acquisition and funding for the FinnDiane Study. AJM acquired and analysed the UK-ROI cohort data and contributed to the interpretation of the study results. NS contributed to FinnDiane data collection, investigation, interpretation of the study results, and funding. AS, CH, LJS, EHD, SM, ZC, RN, SP, AP, HJ, GM, KS, JNH, JCF, APM, P-HG, AJM, and NS reviewed the manuscript draft, and all authors approved the final version.

## Supplementary Figures

**Figure S1.**
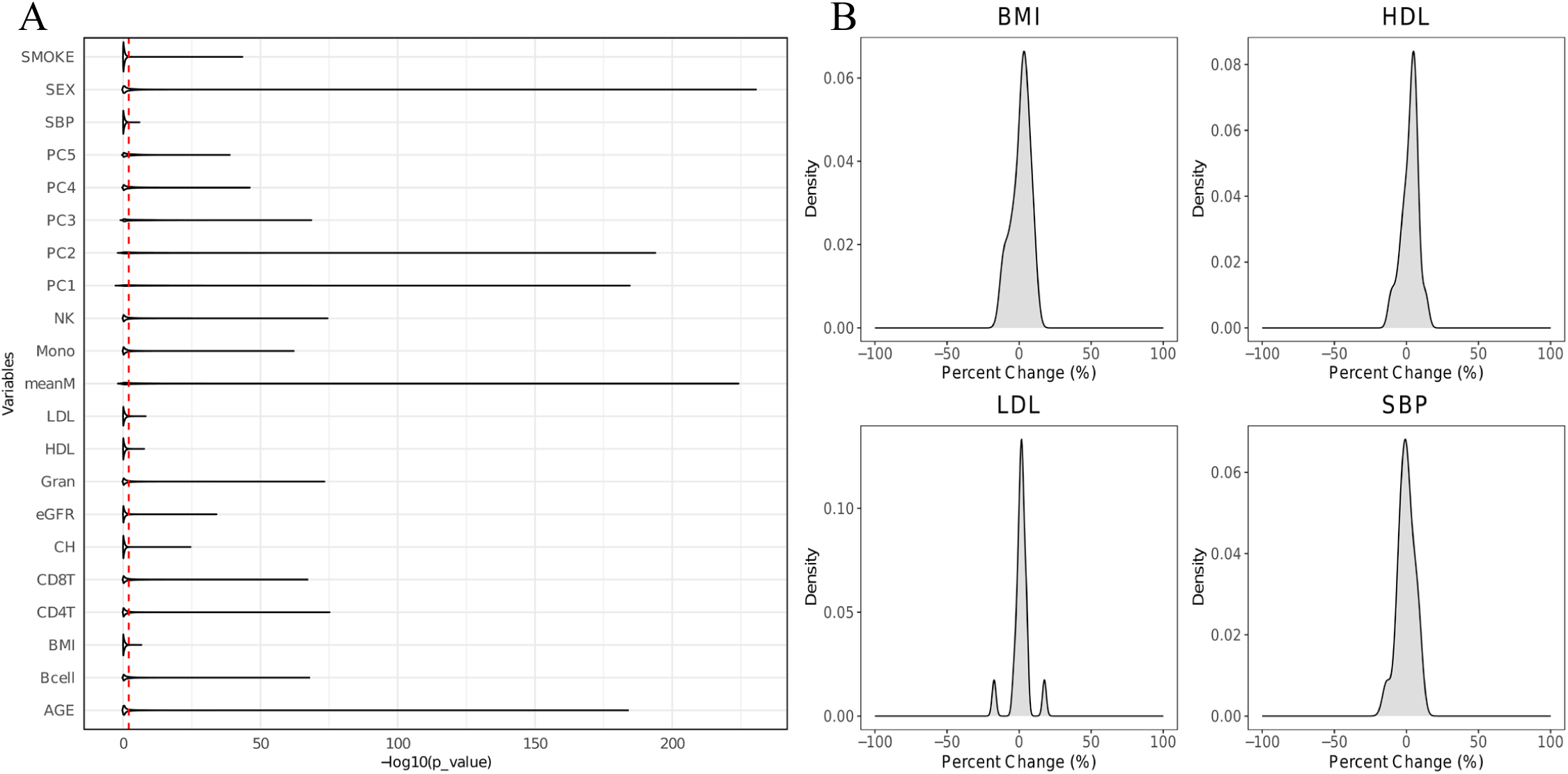
**(A)** Sensitivity analyses. violin plot of association significance between DNA methylation and clinical variables. **(B)** Percentage change distribution of association coefficients between DNA methylation and clinical variables at CH-associated CpGs.

**Figure S2.**
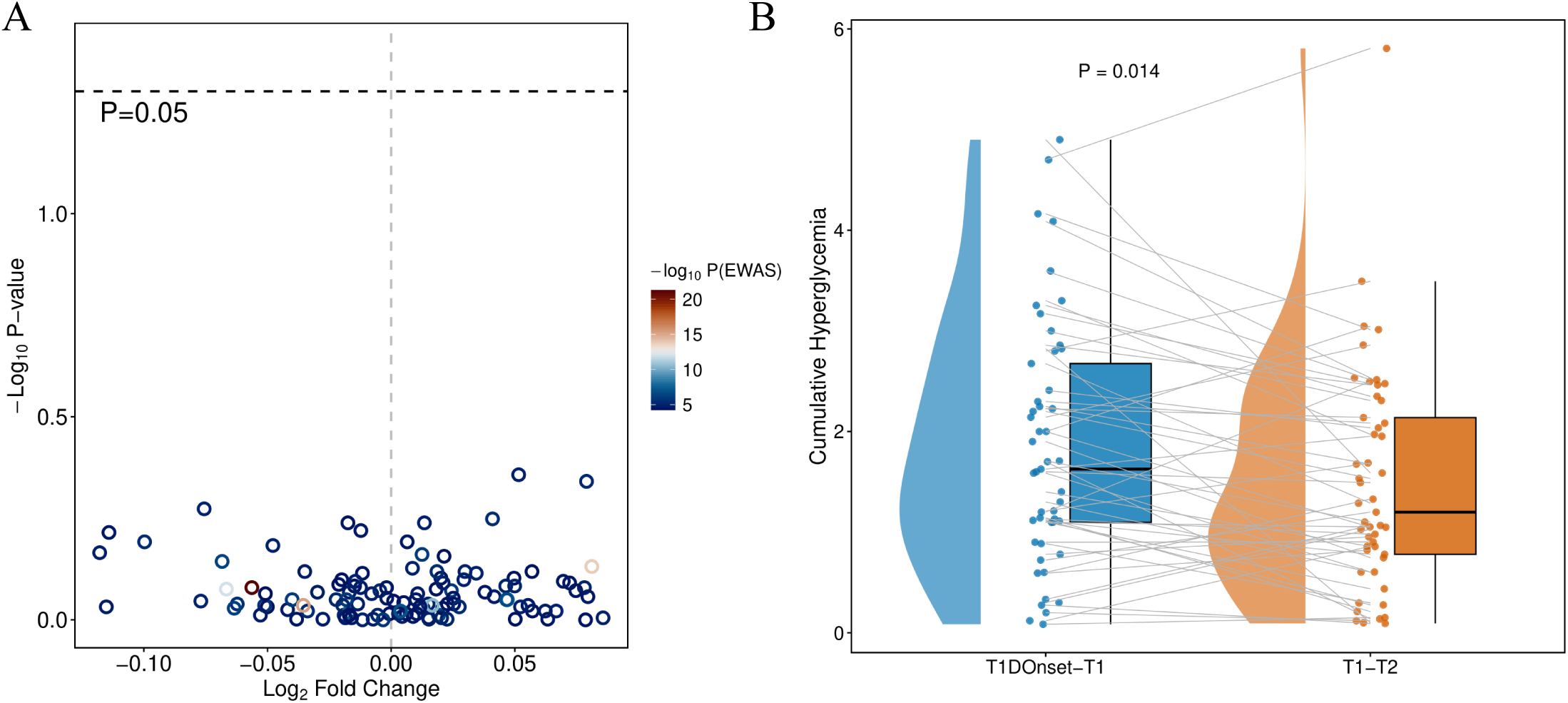
Changes in DNAmet over time at suggestive CH-associated CpGs. **(A)** Volcano plot of changes in DNA methylation at 115 suggestive CH-associated CpGs across two time points (mean follow-up 7.5 years). **(B)** Cumulative hyperglycaemia calculated for the interval from T1D onset to first DNA methylation and from first DNA methylation to second DNA methylation.

**Figure S3.**
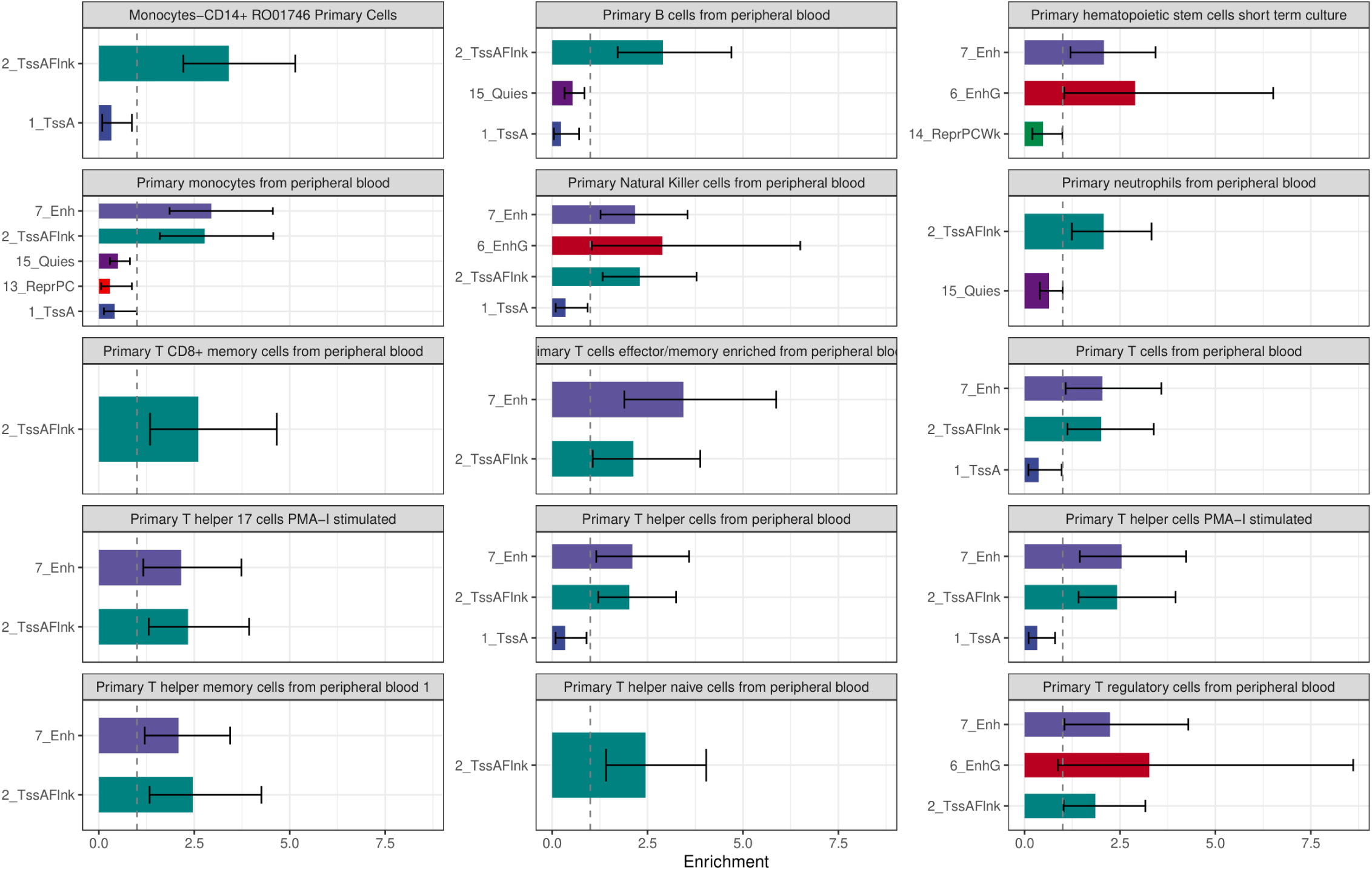
Enrichment of suggestive CH-associated CpGs across ChromHMM states in major blood cell types. Only significant (p<0.05) enrichment results are shown, altogether 27 blood cell types were investigated. Odds ratios (ORs) and 95% confidence intervals were estimated using Fisher’s exact test. Chromatin states are defined according to the 15-state ChromHMM model: active TSS (1_TssA), 2_TssAFlnk (Flanking Activate TSS), 6_EnhG (Genic enhancers), 7_Enh (Enhancers), 13_ReprPC (Repressed PolyComb), 14_ReprPCWk (Weak Repressed Polycomb), 15_Quies (Quiescent/Low).

**Figure S4.**
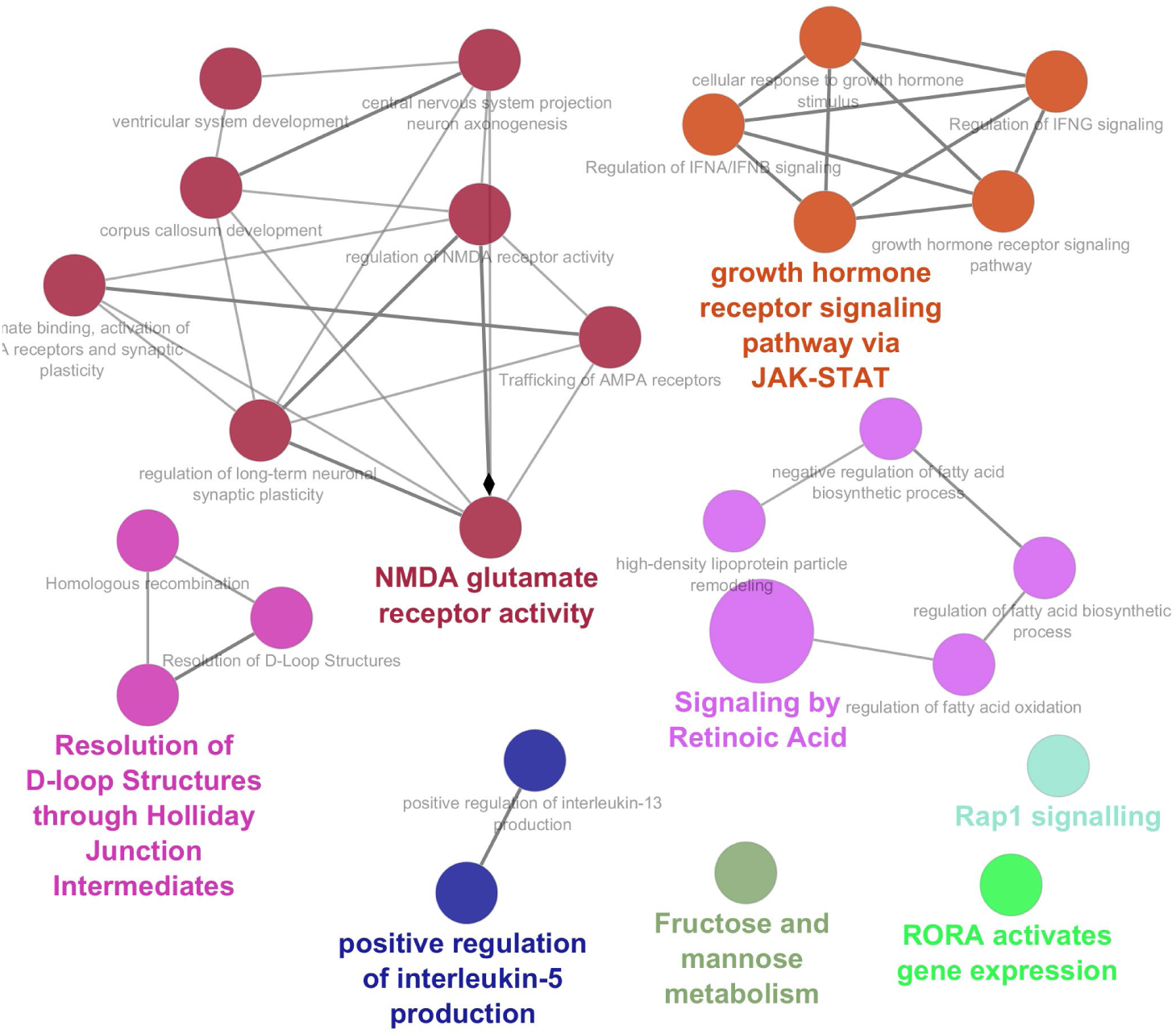
Interactive network of GO and KEGG terms enriched in suggestive CH-associated CpG closest genes generated by the Cytoscape plugin ClueGO.

**Figure S5.**
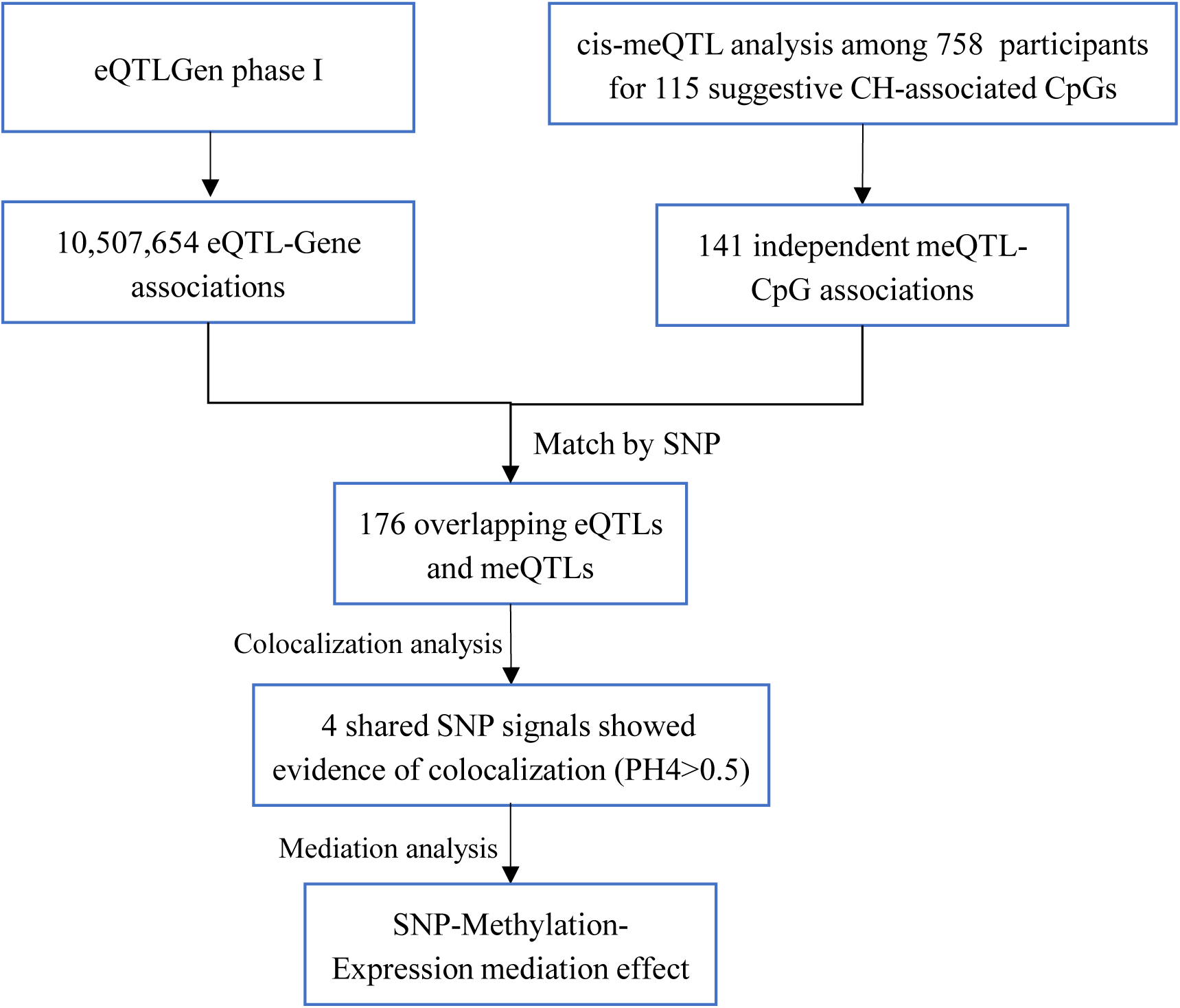
Workflow for identifying SNPs with Colocalisation Effect. 115 suggestive CH-associated CpGs were associated with 141 independent *cis*-meQTLs in 758 FinnDiane participants. 3,663,468 primary *cis*-eQTLs from eQTLGen phase I were overlapped with 68 meQTL signals. Four shared SNP signals showed evidence of colocalisation and were further investigated for SNP-methylation-expression mediation effects.

**Figure S6.**
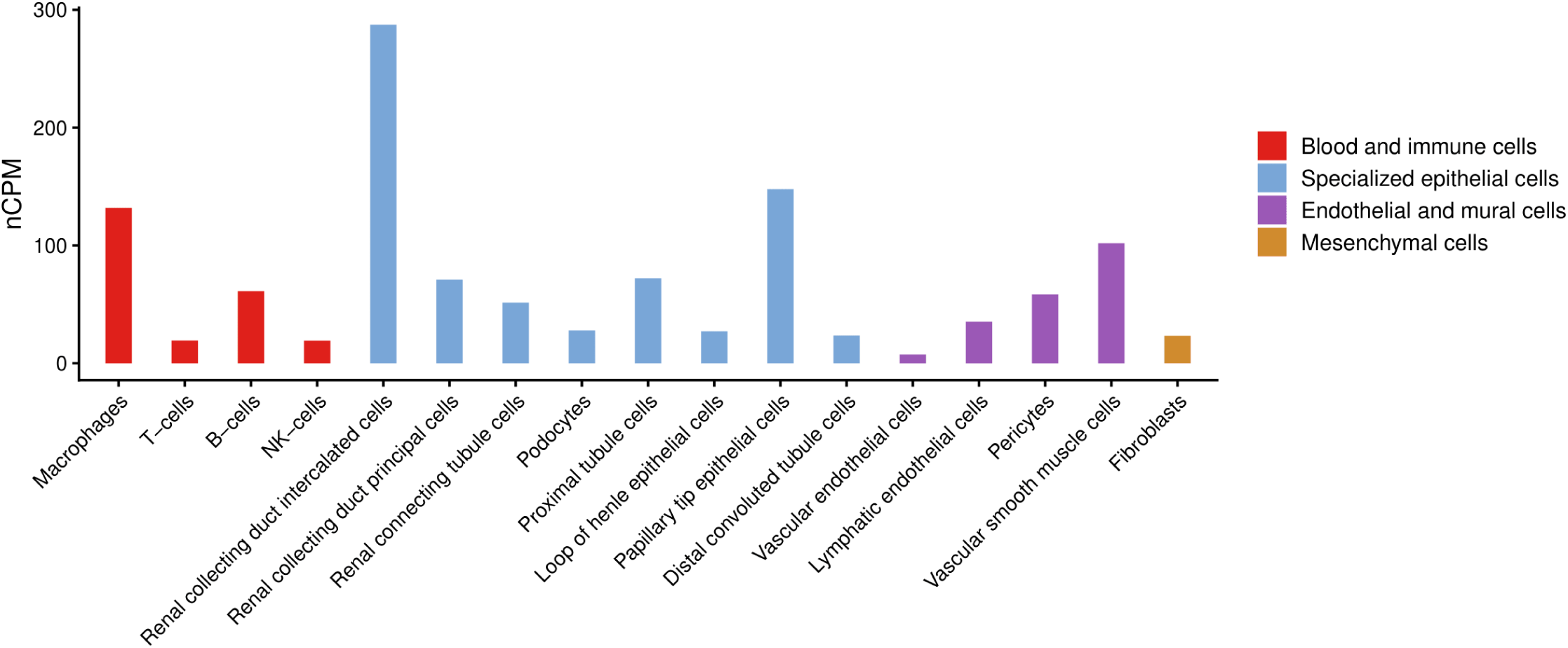
Single-cell expression of PPFIBP2 in kidney. nCPM. normalized counts per million, adapted from https://www.proteinatlas.org/ENSG00000166387-PPFIBP2/single+cell/kidney.

**Figure S7.**
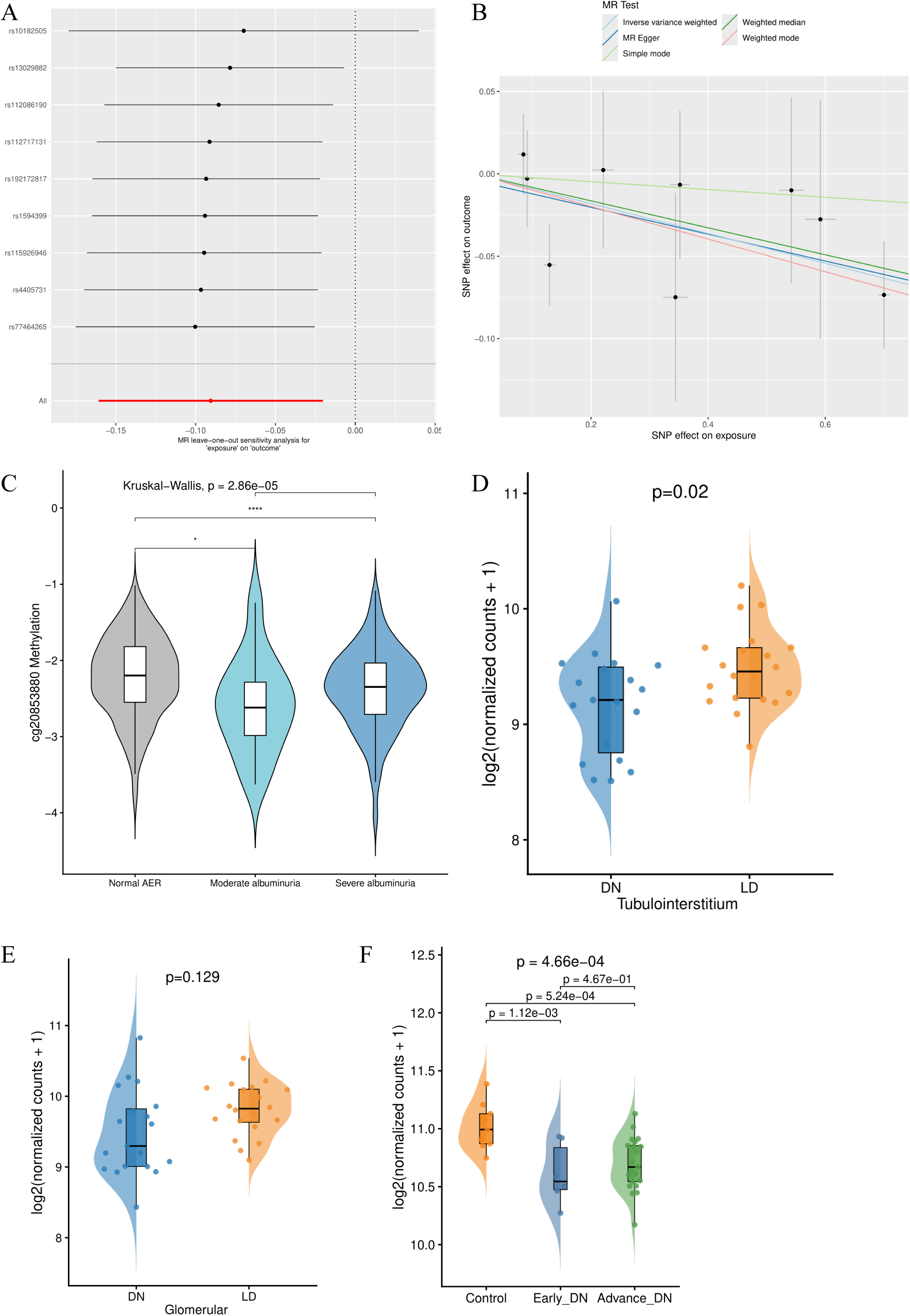
Mendelian Randomization for cg20853880 (*KLF11*) and DKD. **(A)** Leave-one-out sensitivity analysis for MR estimated for the effect of cg20853880 on DKD. Each point represents the MR estimate for the effect of the exposure on the outcome when each SNP is individually excluded from the analysis, and horizontal bars indicate 95% confidence intervals. **(B)** Scatter plot of the relationship of the SNP effects on the exposure against the SNP effects on the outcome. Each point represents a SNP and the slope of fitted line corresponding to the estimated causal effect under different methods. **(C)** Violin plot of cg20853880 methylation levels in participants with normal AER, moderate albuminuria, and severe albuminuria. Overall difference was assessed using the Kruskal–Wallis test. Pairwise comparisons were performed using the Wilcoxon rank-sum test. **(D)** Violin plot of *KLF11* gene expression levels in the tubulointerstitium comparing diabetic nephropathy (DN, n=19) and living donor (LD, n=20) samples based on re-analysis of data from *Levin et al*^82^. Expression was transformed as log_2_(normalized count + 1). **(E)** Violin plot of *KLF11* gene expression levels in glomerular comparing diabetic nephropathy (DN, n=19) and living donor (LD, n=20) samples. Expression was transformed as log_2_(normalized count + 1). **(F)** Violin plot of *KLF11* gene expression levels in control human kidney samples (Control, n=9), early diabetic nephropathy (Early_DN, n=6), and advanced diabetic nephropathy (Advance_DN, n = 21).

**Figure S8.**
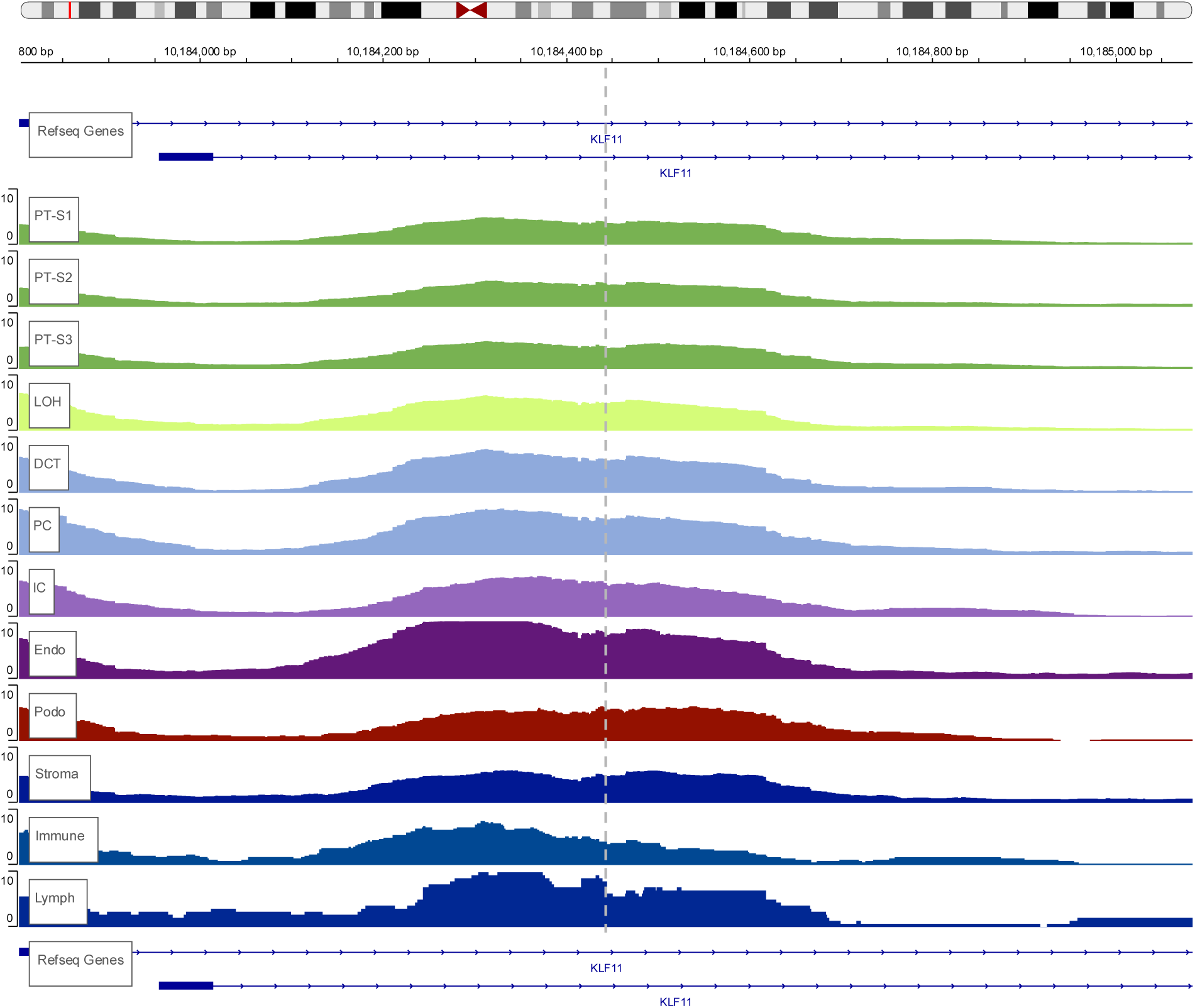
Open chromatin peaks in kidney cell types. Figure is adapted from https://susztaklab.com/Human_snATAC/index.php (chr2:10,183,800-10,185,050, hg19), and cg20853880 (chr2:10184444, hg19) position is incorporated. Each track represents a cell type: proximal tubule S1-S3 segments (PT-S1-PT-S3), loop of Henle (LOH), distal convoluted tubule (DCT), principal cell of collecting duct (PC), intercalated cell of collecting duct (IC), endothelial cells (Endo), podocyte (Podo), stroma cells (Stroma), immune cells (Immune), lymph cells (Lymph).

**Figure S9.**
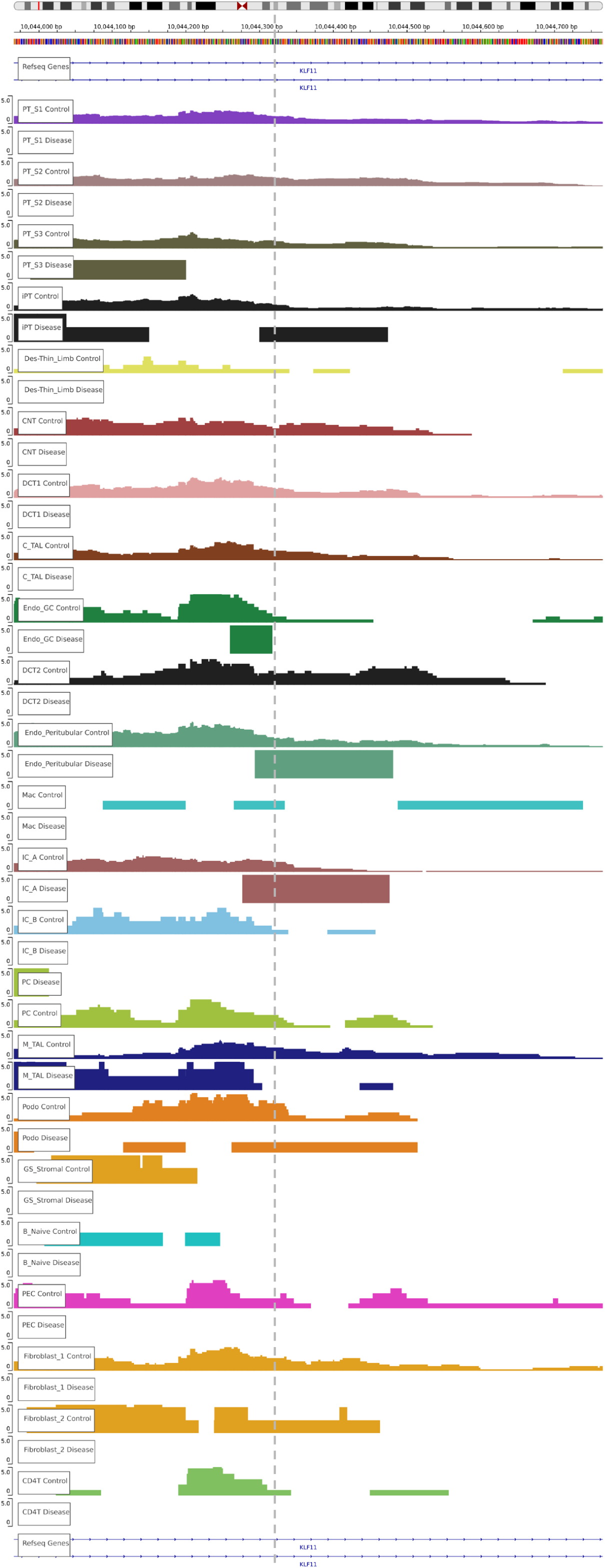
The full-sized track of Figure 5D.

**Figure S10.**
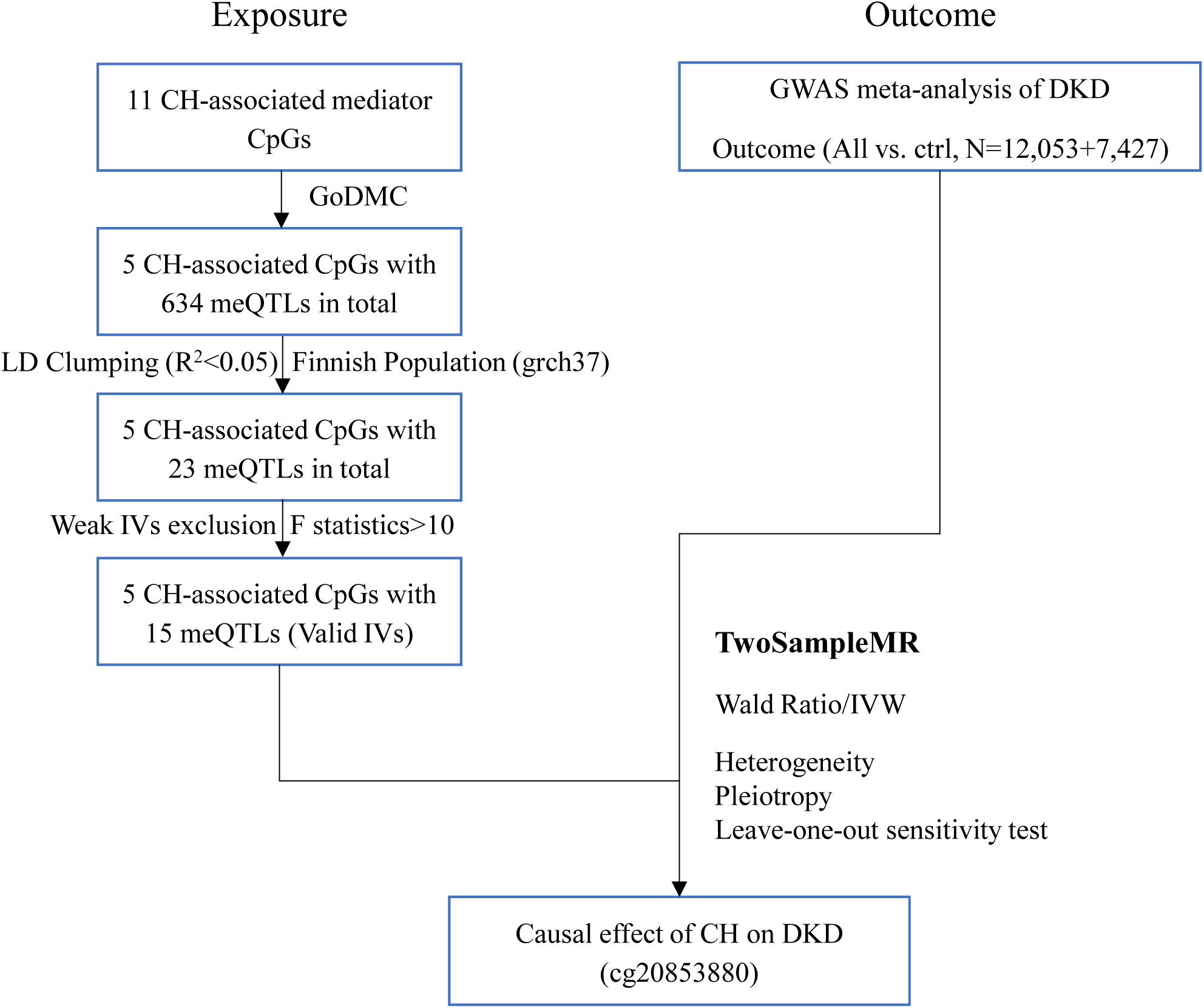
Workflow of TwoSampleMR of CH-associated CpGs on DKD risk. Among. **s**eventeen CH-associated CpGs, 8 of them had a total of 966 *cis*-meQTLs, and 42 independent *cis*-meQTLs were retained after LD Clumping. 25 valid instrumental variables for 8 CH-associated CpGs were kept after weak instrumental variables exclusion. Summary statistics of DKD were obtained from GWAS meta-analysis of DKD as outcome. Heterogeneity, pleiotropy, and leave-one-out sensitivity test were performed.

## Extended Data Figure

**Extended Data Fig.1.**
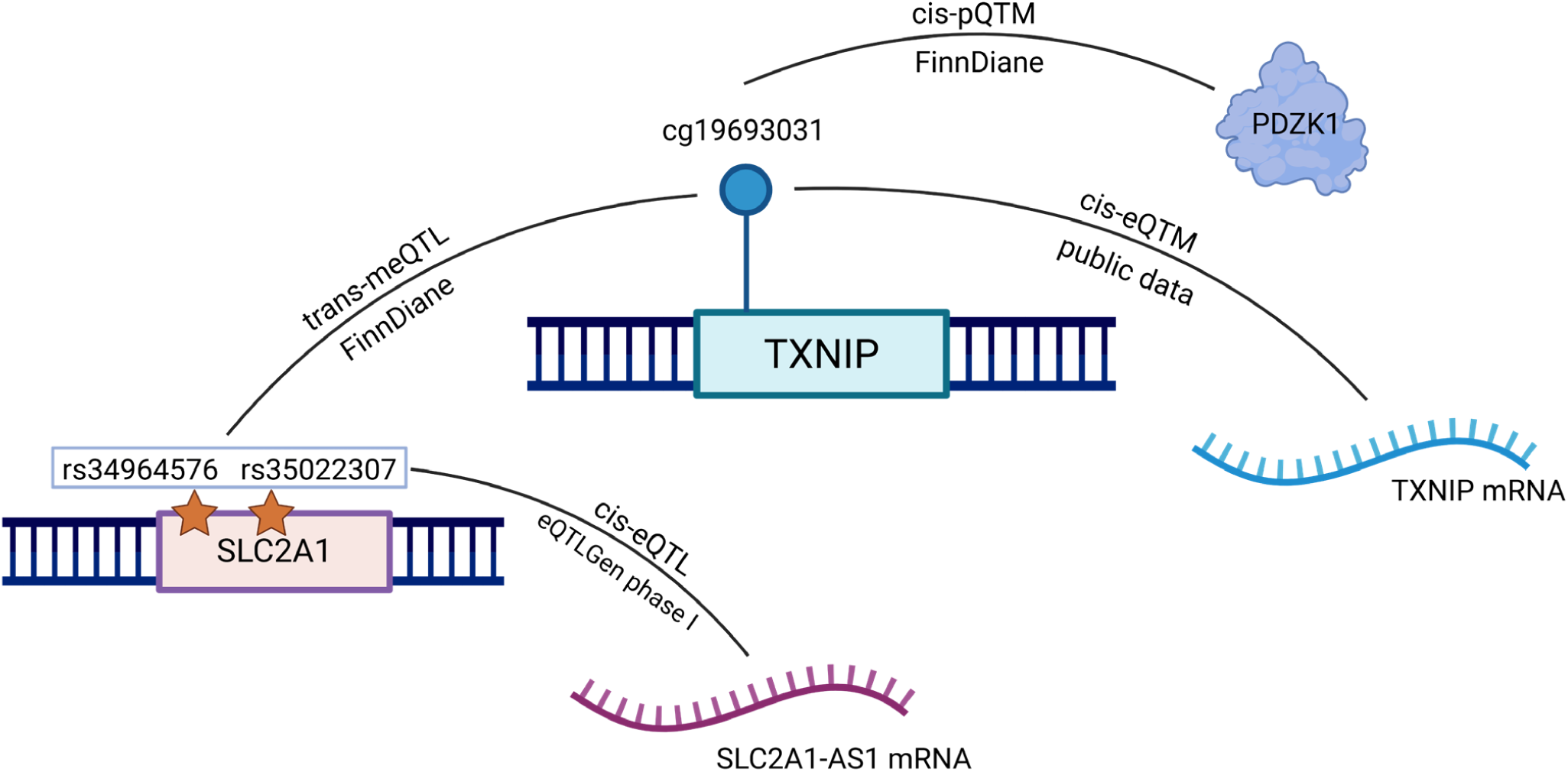
Integrative Multi-omics Associations of the cg19693031 (*TXNIP*) CpG Site. The most significant CH-associated CpG (cg19693031 at 3’UTR of TXNIP) was associated with serum PDZK1 protein level in FinnDiane proteomics data (N=292). cg19693031 was also associated with two trans-meQTL variants, rs35022307 and rs34964576 in *SLC2A1,* in FinnDiane (N=758). Both variants were associated with *SLC2A-AS1* (*SLC2A1* divergent transcript) expression^26^. cg19693031 was associated with *TXNIP* mRNA expression level in kidney^20^, monocytes^23^, and whole blood^21,22^. Created in BioRender. Luo, X. (2026) https://BioRender.com/n08w0rl

**Extended Data Fig. 2.**
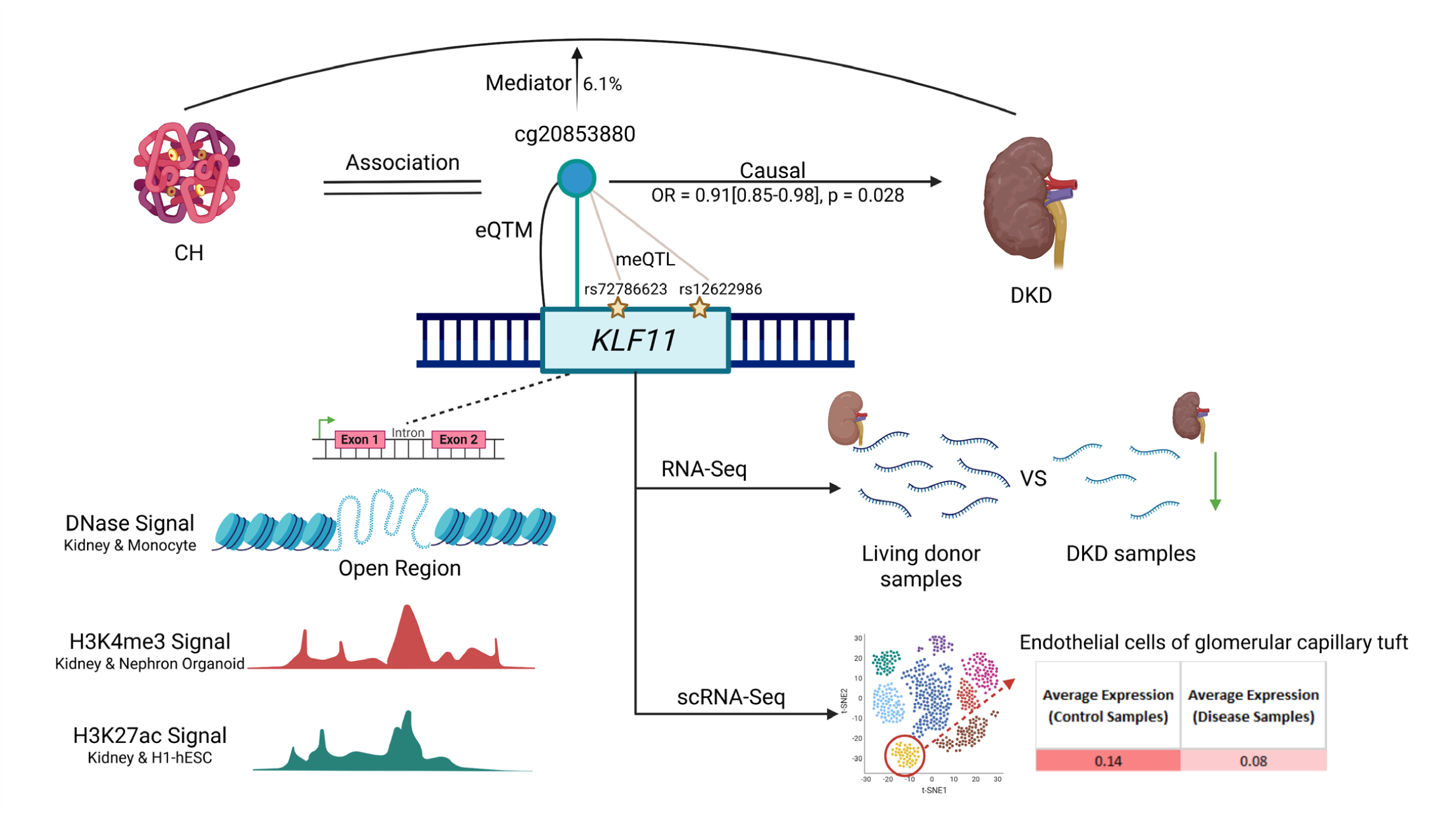
Integrated evidence supporting *KLF11*/cg20853880 as a candidate regulatory locus linking cumulative hyperglycaemia to DKD. Created in BioRender. Luo, X. (2026) https://BioRender.com/mdu4yauff

## References

1. Magliano, D. & Boyko, E. J. Diabetes Atlas. (International Diabetes Federation, Brussels, 2025).

2. Ogle, G. D. et al. Global type 1 diabetes prevalence, incidence, and mortality estimates 2025: Results from the International diabetes Federation Atlas, 11th Edition, and the T1D Index Version 3.0. Diabetes Research and Clinical Practice 225, 112277 (2025).

3. Heerspink, H. J. et al. People with type 1 diabetes and chronic kidney disease urgently need new therapies: a call for action. Lancet Diabetes Endocrinol 11, 536–540 (2023).

4. Diabetes Control and Complications Trial/Epidemiology of Diabetes Interventions and Complications Research Group et al. Retinopathy and nephropathy in patients with type 1 diabetes four years after a trial of intensive therapy. N Engl J Med 342, 381–389 (2000).

5. Lachin, J. M., Nathan, D. M., & DCCT/EDIC Research Group. Understanding Metabolic Memory: The Prolonged Influence of Glycemia During the Diabetes Control and Complications Trial (DCCT) on Future Risks of Complications During the Study of the Epidemiology of Diabetes Interventions and Complications (EDIC). Diabetes Care 44, 2216–2224 (2021).

6. Aranyi, T. & Susztak, K. Cytosine Methylation Studies in Patients with Diabetic Kidney Disease. Curr Diab Rep 19, 91 (2019).

7. Park, L. K. et al. Genome-wide DNA methylation analysis identifies a metabolic memory profile in patient-derived diabetic foot ulcer fibroblasts. Epigenetics 9, 1339–1349 (2014).

8. Chen, Z. et al. DNA methylation mediates development of HbA1c-associated complications in type 1 diabetes. Nat Metab 2, 744–762 (2020).

9. Guo, J. et al. Accelerated Kidney Aging in Diabetes Mellitus. Oxidative Medicine and Cellular Longevity 2020, 1–24 (2020).

10. Pan, Y. et al. Effects of epigenetic age acceleration on kidney function: a Mendelian randomization study. Clin Epigenet 15, 61 (2023).

11. Sheng, X. et al. Systematic integrated analysis of genetic and epigenetic variation in diabetic kidney disease. Proc. Natl. Acad. Sci. U.S.A. 117, 29013–29024 (2020).

12. Nuotio, M.-L. et al. An epigenome-wide association study of metabolic syndrome and its components. Sci Rep 10, 20567 (2020).

13. Cataldi, S., Costa, V., Ciccodicola, A. & Aprile, M. PPARγ and Diabetes: Beyond the Genome and Towards Personalized Medicine. Curr Diab Rep 21, 18 (2021).

14. Parmar, U. M., Jalgaonkar, M. P., Kansara, A. J. & Oza, M. J. Emerging links between FOXOs and diabetic complications. European Journal of Pharmacology 960, 176089 (2023).

15. Yang, F. et al. EWAS Open Platform 2026: a deeply integrated resource for epigenome-wide association studies. Nucleic Acids Res 54, D1061–D1068 (2026).

16. Ernst, J. & Kellis, M. ChromHMM: automating chromatin-state discovery and characterization. Nat Methods 9, 215–216 (2012).

17. Min, J. L. et al. Genomic and phenotypic insights from an atlas of genetic effects on DNA methylation. Nat Genet 53, 1311–1321 (2021).

18. Villicaña, S. et al. Genetic impacts on DNA methylation help elucidate regulatory genomic processes. Genome Biol 24, 176 (2023).

19. Miller, R. G. et al. DNA methylation and 28-year cardiovascular disease risk in type 1 diabetes: the Epidemiology of Diabetes Complications (EDC) cohort study. Clinical Epigenetics (2023).

20. Liu, H. et al. Epigenomic and transcriptomic analyses define core cell types, genes and targetable mechanisms for kidney disease. Nat Genet 54, 950–962 (2022).

21. Ruiz-Arenas, C. et al. Identification of autosomal cis expression quantitative trait methylation (cis eQTMs) in children’s blood. eLife 11, e65310 (2022).

22. the BIOS Consortium et al. Disease variants alter transcription factor levels and methylation of their binding sites. Nat Genet 49, 131–138 (2017).

23. Kennedy, E. M. et al. An integrated-omics analysis of the epigenetic landscape of gene expression in human blood cells. BMC Genomics 19, 476 (2018).

24. Xie, Y. et al. CPT1A Protects Podocytes From Lipotoxicity and Apoptosis In Vitro and Alleviates Diabetic Nephropathy In Vivo. Diabetes 73, 879–895 (2024).

25. Garcia Galindo, J. J., et al. Association of Netrin 1 with hsCRP in Subjects with Obesity and Recent Diagnosis of Type 2 Diabetes. Curr Issues Mol Biol 45, 134–140 (2022).

26. Võsa, U. et al. Large-scale cis-and trans-eQTL analyses identify thousands of genetic loci and polygenic scores that regulate blood gene expression. Nat Genet 53, 1300–1310 (2021).

27. Salem, R. M. et al. Genome-Wide Association Study of Diabetic Kidney Disease Highlights Biology Involved in Glomerular Basement Membrane Collagen. J Am Soc Nephrol 30, 2000–2016 (2019).

28. Yan, Y. et al. Unraveling the epigenetic code: human kidney DNA methylation and chromatin dynamics in renal disease development. Nat Commun 15, 873 (2024).

29. Fan, Y. et al. Comparison of Kidney Transcriptomic Profiles of Early and Advanced Diabetic Nephropathy Reveals Potential New Mechanisms for Disease Progression. Diabetes 68, 2301–2314 (2019).

30. Abedini, A. et al. Single-cell multi-omic and spatial profiling of human kidneys implicates the fibrotic microenvironment in kidney disease progression. Nat Genet 56, 1712–1724 (2024).

31. Miller, R. G., Mychaleckyj, J. C., Onengut-Gumuscu, S., Orchard, T. J. & Costacou, T. TXNIP DNA methylation is associated with glycemic control over 28 years in type 1 diabetes: findings from the Pittsburgh Epidemiology of Diabetes Complications (EDC) study. BMJ Open Diabetes Res Care 11, e003068 (2023).

32. Kulkarni, H. et al. Novel epigenetic determinants of type 2 diabetes in Mexican-American families. Hum Mol Genet 24, 5330–5344 (2015).

33. Xiang, Y. et al. DNA Methylation of TXNIP Independently Associated with Inflammation and Diabetes Mellitus in Twins. Twin Res Hum Genet 24, 273–280 (2021).

34. Smyth, L. J. et al. Epigenome-wide meta-analysis identifies DNA methylation biomarkers associated with diabetic kidney disease. Nat Commun 13, 7891 (2022).

35. Das, M. et al. Association of DNA Methylation at CPT1A Locus with Metabolic Syndrome in the Genetics of Lipid Lowering Drugs and Diet Network (GOLDN) Study. PLoS One 11, e0145789 (2016).

36. Bibikova, M. et al. High density DNA methylation array with single CpG site resolution. Genomics 98, 288–295 (2011).

37. Belsky, D. W. et al. DunedinPACE, a DNA methylation biomarker of the pace of aging. Elife 11, e73420 (2022).

38. Sarkar, A., Saha, S. & Bhattacharya, D. The effect of chronic hyperglycemia on aging: implications in cognitive impairment. in Diabetes and Neurodegeneration 237–251 (Elsevier, 2026). doi:10.1016/B978-0-443-33355-2.00027-X.

39. Meissner, C. & Ritz-Timme, S. Molecular pathology and age estimation. Forensic Sci Int 203, 34–43 (2010).

40. Yamagishi, S.-I., Nakamura, N., Suematsu, M., Kaseda, K. & Matsui, T. Advanced Glycation End Products: A Molecular Target for Vascular Complications in Diabetes. Mol Med 21 Suppl 1, S32–40 (2015).

41. Adeshara, K. et al. Protein glycation products associate with progression of kidney disease and incident cardiovascular events in individuals with type 1 diabetes. Cardiovasc Diabetol 23, 235 (2024).

42. The Human Protein Atlas https://www.proteinatlas.org/ENSG00000174827-PDZK1/single+cell/kidney.

43. Lu, S. et al. Downregulation of PDZK1 by TGF-β1 promotes renal fibrosis via inducing epithelial-mesenchymal transition of renal tubular cells. Biochem Pharmacol 220, 116015 (2024).

44. Zhao, J. et al. PDZK1 Protects Against RPE Senescence by Targeting the 14-3-3ε-mTOR Axis to Attenuate Early Diabetic Retinopathy. Adv Sci (Weinh) 12, e11288 (2025).

45. Junyent, M. et al. Genetic variants at the PDZ-interacting domain of the scavenger receptor class B type I interact with diet to influence the risk of metabolic syndrome in obese men and women. J Nutr 139, 842–848 (2009).

46. Syreeni, A. et al. Blood DNA methylation markers are associated with diabetic kidney disease progression in type 1 diabetes. Diabetologia (2026) doi:10.1007/s00125-025-06661-7.

47. Ryuge, A., et al. Basigin deficiency prevents anaplerosis and ameliorates insulin resistance and hepatosteatosis. JCI Insight 6, e142464 (2021).

48. Wang, X., Shi, L., Han, Z. & Liu, B. Follistatin-like 3 suppresses cell proliferation and fibronectin expression via p38MAPK pathway in rat mesangial cells cultured under high glucose. Int J Clin Exp Med 8, 15214–15221 (2015).

49. Hua, Y., Yin, Z., Li, M., Sun, H. & Shi, B. Correlation between circulating advanced glycation end products and thioredoxin-interacting protein levels and renal fat content in type 2 diabetes mellitus patients. Diabetol Metab Syndr 16, 144 (2024).

50. Wang, L., Zhu, B., Liu, S., Xingmei, Y. & Zhou, Y. Blocking Intracellular TXNIP Shuttling Attenuates Reactive Oxygen Species-Associated NLRP3 Inflammasome Activation Induced by Tubulointerstitial Fibrosis in Diabetic Kidney Disease: SA-PO256. Journal of the American Society of Nephrology 35, (2024).

51. De Marinis, Y. et al. Epigenetic regulation of the thioredoxin-interacting protein (TXNIP) gene by hyperglycemia in kidney. Kidney International 89, 342–353 (2016).

52. Lomberk, G. et al. Krüppel-like Factor 11 Regulates the Expression of Metabolic Genes via an Evolutionarily Conserved Protein Interaction Domain Functionally Disrupted in Maturity Onset Diabetes of the Young. Journal of Biological Chemistry 288, 17745–17758 (2013).

53. Park, S. G., Hannenhalli, S. & Choi, S. S. Conservation in first introns is positively associated with the number of exons within genes and the presence of regulatory epigenetic signals. BMC Genomics 15, 526 (2014).

54. Malekkou, A. et al. A novel mutation deep within intron 7 of the *GBA* gene causes Gaucher disease. Molec Gen & Gen Med 8, e1090 (2020).

55. Ong, C.-T. & Adusumalli, S. Increased intron retention is linked to Alzheimer’s disease. Neural Regen Res 15, 259 (2020).

56. Neve, B. et al. Role of transcription factor KLF11 and its diabetes-associated gene variants in pancreatic beta cell function. Proc Natl Acad Sci U S A 102, 4807–4812 (2005).

57. Zhang, H. et al. Mouse KLF11 regulates hepatic lipid metabolism. J Hepatol 58, 763–770 (2013).

58. Zhang, H. et al. Mouse KLF11 regulates hepatic lipid metabolism. J Hepatol 58, 763–770 (2013).

59. Niu, X. et al. Human Krüppel-like factor 11 inhibits human proinsulin promoter activity in pancreatic beta cells. Diabetologia 50, 1433–1441 (2007).

60. Rane, M. J., Zhao, Y. & Cai, L. Krϋppel-like factors (KLFs) in renal physiology and disease. EBioMedicine 40, 743–750 (2019).

61. Nath, K. A. et al. KLF11 Is a Novel Endogenous Protectant against Renal Ischemia-Reperfusion Injury. Kidne*y360* 3, 1417–1422 (2022).

62. De Lorenzo, S. B. et al. KLF11 deficiency enhances chemokine generation and fibrosis in murine unilateral ureteral obstruction. PLoS One 17, e0266454 (2022).

63. Wu, W., Wang, X., Yu, X. & Lan, H.-Y. Smad3 Signatures in Renal Inflammation and Fibrosis. Int J Biol Sci 18, 2795–2806 (2022).

64. Matsumori, A. Nuclear Factor-κB is a Prime Candidate for the Diagnosis and Control of Inflammatory Cardiovascular Disease. Eur Cardiol 18, e40 (2023).

65. Tang, J., Liu, F., Cooper, M. E. & Chai, Z. Renal fibrosis as a hallmark of diabetic kidney disease: potential role of targeting transforming growth factor-beta (TGF-β) and related molecules. Expert Opin Ther Targets 26, 721–738 (2022).

66. Foresto-Neto, O., et al. SUN-311 NF-KAPPA B ACTIVATION PROMOTES GLOMERULAR INJURY AND INFLAMMATION IN LONG-TERM EXPERIMENTAL DIABETIC KIDNEY DISEASE. Kidney International Reports 4, S289 (2019).

67. Yoshida, T., Yamashita, M., Iwai, M. & Hayashi, M. Endothelial Krüppel-Like Factor 4 Mediates the Protective Effect of Statins against Ischemic AKI. J Am Soc Nephrol 27, 1379–1388 (2016).

68. Parmar, K. M. et al. Statins exert endothelial atheroprotective effects via the KLF2 transcription factor. J Biol Chem 280, 26714–26719 (2005).

69. Sen-Banerjee, S. et al. Kruppel-like factor 2 as a novel mediator of statin effects in endothelial cells. Circulation 112, 720–726 (2005).

70. Tuomisto, T. T. et al. Simvastatin has an anti-inflammatory effect on macrophages via upregulation of an atheroprotective transcription factor, Kruppel-like factor 2. Cardiovasc Res 78, 175–184 (2008).

71. Xu, Y. et al. Tannic acid as a plant-derived polyphenol exerts vasoprotection via enhancing KLF2 expression in endothelial cells. Sci Rep 7, 6686 (2017).

72. Gracia-Sancho, J., Villarreal, G., Zhang, Y. & García-Cardeña, G. Activation of SIRT1 by resveratrol induces KLF2 expression conferring an endothelial vasoprotective phenotype. Cardiovasc Res 85, 514–519 (2010).

73. Grunewald, M. et al. Mechanistic role for a novel glucocorticoid-KLF11 (TIEG2) protein pathway in stress-induced monoamine oxidase A expression. J Biol Chem 287, 24195–24206 (2012).

74. Chu, A. Y. et al. Epigenome-wide association studies identify DNA methylation associated with kidney function. Nat Commun 8, 1286 (2017).

75. Edgar, R. D., Jones, M. J., Robinson, W. P. & Kobor, M. S. An empirically driven data reduction method on the human 450K methylation array to remove tissue specific non-variable CpGs. Clin Epigenetics 9, 11 (2017).

76. Houseman, E. A. et al. DNA methylation arrays as surrogate measures of cell mixture distribution. BMC Bioinformatics 13, 86 (2012).

77. Inker, L. A. et al. New Creatinine-and Cystatin C-Based Equations to Estimate GFR without Race. N Engl J Med 385, 1737–1749 (2021).

78. Chen, B. H. & Zhou, W. mLiftOver: harmonizing data across Infinium DNA methylation platforms. Bioinformatics 40, btae423 (2024).

79. Hill, C. et al. Baseline epigenetic clock measures in the Northern Ireland Cohort for the Longitudinal Study of Ageing (NICOLA). BMC Res Notes 19, 192 (2026).

80. Higgins-Chen, A. T. et al. A computational solution for bolstering reliability of epigenetic clocks: implications for clinical trials and longitudinal tracking. Nat Aging 2, 644–661 (2022).

81. Sandholm, N. et al. Whole-exome sequencing identifies novel protein-altering variants associated with serum apolipoprotein and lipid concentrations. Genome Med 14, 132 (2022).

82. Levin, A. et al. Novel insights into the disease transcriptome of human diabetic glomeruli and tubulointerstitium. Nephrol Dial Transplant 35, 2059–2072 (2020).

83. Anders, S. & Huber, W. Differential expression analysis for sequence count data. Genome Biol 11, R106 (2010).

84. Fan, Y. et al. Erratum. Comparison of Kidney Transcriptomic Profiles of Early and Advanced Diabetic Nephropathy Reveals Potential New Mechanisms for Disease Progression. Diabetes 2019;68:2301-2314. Diabetes 69, 797 (2020).

